# Serological reconstruction of COVID-19 epidemics through analysis of antibody kinetics to SARS-CoV-2 proteins

**DOI:** 10.1101/2021.03.04.21252532

**Authors:** Stéphane Pelleau, Tom Woudenberg, Jason Rosado, Françoise Donnadieu, Laura Garcia, Thomas Obadia, Soazic Gardais, Yasmine Elgharbawy, Aurelie Velay, Maria Gonzalez, Jacques Yves Nizou, Nizar Khelil, Konstantinos Zannis, Charlotte Cockram, Sarah Hélène Merkling, Annalisa Meola, Solen Kerneis, Benjamin Terrier, Jerome de Seze, Delphine Planas, Olivier Schwartz, François Dejardin, Stéphane Petres, Cassandre von Platen, Laurence Arowas, Louise Perrin de Facci, Darragh Duffy, Clíona Ní Cheallaigh, Niall Conlon, Liam Townsend, Heidi Auerswald, Marija Backovic, Bruno Hoen, Arnaud Fontanet, Ivo Mueller, Samira Fafi-Kremer, Timothée Bruel, Michael White

**Affiliations:** Malaria: Parasites and Hosts Unit, Department of Parasites and Insect Vectors, Institut Pasteur, Paris, France; Sorbonne Université, ED 393, F-75005 Paris, France; Hub de Bioinformatique et Biostatistique, Département Biologie Computationnelle, Institut Pasteur, Paris, France; CHU de Strasbourg, Laboratoire de Virologie, F-67091 Strasbourg, France; Université de Strasbourg, INSERM, IRM UMR_S 1109, Strasbourg, France; CHU de Strasbourg, Service de Pathologies Professionnelles, F-67091 Strasbourg, France; Institut Mutualiste Montsouris, Paris, France; Spatial Regulation of Genomes Unit, Department of Genomes and Genetics, Institut Pasteur, Paris, France; Insect-Virus Interactions Unit, Department of Virology and CNRS UMR 2000, Institut Pasteur, Paris, France; Structural Virology Unit, Department of Virology and CNRS UMR 3569, Institut Pasteur, Paris, France; Equipe Mobile d’Infectiologie, APHP Centre-Université de Paris, Paris, France; Epidemiology and Modelling of Bacterial Escape to Antimicrobials Unit, Department of Global Health, Institut Pasteur, Paris, France; Department of Internal Medicine, National Referral Center for Rare Systemic Autoimmune Diseases, Assistance Publique Hôpitaux de Paris-Centre (APHP-CUP), Université de Paris, Paris, France; PARCC, INSERM U970, Paris, France; Centre d’Investigation Clinique - INSERM CIC-1434, Strasbourg, France; Virus and Immunity Unit, Department of Virology, Institut Pasteur, Paris, France; Production and Purification of Recombinant Proteins Technological Platform, Center for Technological Resources and Research, Institut Pasteur, Paris, France; Center for Translational Science, Institut Pasteur, Paris, France; Investigation Clinique et Accès aux Ressources Biologiques (ICAReB), Center for Translational Research, Institut Pasteur, Paris, France; Translational Immunology Lab, Institut Pasteur, Paris, France; Department of Infectious Diseases, St James’s Hospital, Dublin, Ireland; Department of Clinical Medicine, School of Medicine, Trinity Translational Medicine Institute, Trinity College Dublin, Ireland; Department of Immunology, St James’s Hospital, Dublin, Ireland; Department of Immunology, School of Medicine, Trinity College Dublin, Ireland; Virology Unit, Institut Pasteur du Cambodge, Institut Pasteur International Network, Phnom Penh, Cambodia; Direction de la Recherche Médicale, Institut Pasteur, Paris, France; Epidemiology of Emerging Diseases Unit, Department of Global Health, Institut Pasteur, Paris, France; Conservatoire National des Arts et Métiers, Paris, France; Division of Population Health and Immunity, The Walter and Eliza Hall Institute, Melbourne, Australia; Department of Medical Biology, University of Melbourne, Melbourne, Australia; Vaccine Research Institute, Creteil, France

## Abstract

Infection with severe acute respiratory syndrome coronavirus 2 (SARS-CoV-2) induces a complex antibody response that varies by orders of magnitude between individuals and over time. Waning antibody levels lead to reduced sensitivity of serological diagnostic tests over time. This undermines the utility of serological surveillance as the SARS-CoV-2 pandemic progresses into its second year. Here we develop a multiplex serological test for measuring antibodies of three isotypes (IgG, IgM, IgA) to five SARS-CoV-2 antigens (Spike (S), receptor binding domain (RBD), Nucleocapsid (N), Spike subunit 2, Membrane-Envelope fusion) and the Spike proteins of four seasonal coronaviruses. We measure antibody responses in several cohorts of French and Irish hospitalized patients and healthcare workers followed for up to eleven months after symptom onset. The data are analysed with a mathematical model of antibody kinetics to quantify the duration of antibody responses accounting for inter-individual variation. One year after symptoms, we estimate that 36% (95% range: 11%, 94%) of anti-S IgG remains, 31% (9%, 89%) anti-RBD IgG remains, and 7% (1%, 31%) anti-N IgG remains. Antibodies of the IgM isotype waned more rapidly, with 9% (2%, 32%) anti-RBD IgM remaining after one year. Antibodies of the IgA isotype also waned rapidly, with 10% (3%, 38%) anti-RBD IgA remaining after one year. Quantitative measurements of antibody responses were used to train machine learning algorithms for classification of previous infection and estimation of time since infection. The resulting diagnostic test classified previous infections with 99% specificity and 98% (95% confidence interval: 94%, 99%) sensitivity, with no evidence for declining sensitivity over the time scale considered. The diagnostic test also provided accurate classification of time since infection into intervals of 0 – 3 months, 3 – 6 months, and 6 – 12 months. Finally, we present a computational method for serological reconstruction of past SARS-CoV-2 transmission using the data from this test when applied to samples from a single cross-sectional sero-prevalence survey.

## Introduction

Severe acute respiratory syndrome coronavirus 2 (SARS-CoV-2), causing coronavirus disease 2019 (COVID-19), has led to widespread morbidity and mortality since its emergence. The response to the SARS-CoV-2 pandemic is critically dependent on surveillance data, most notably numbers of COVID-19 associated hospital admissions and deaths recorded through health systems surveillance, as well as numbers of cases confirmed SARS-CoV-2 positive by PCR-based testing^1^. Other tools are providing crucial complementary information, for example genomic surveillance has been key to tracking the emergence of novel SARS-CoV-2 variants^2^. Serology, based on the detection of antibodies induced by previous infection with SARS-CoV-2, represents another category of surveillance information^3,4^. Appropriately designed sero-prevalence studies can provide estimates of the proportion of a population who have been previously infected. Although no substitute for health systems surveillance, sero-prevalence studies have the advantage of accounting for asymptomatic cases, and symptomatic individuals who do not present to health systems. Sero-prevalence studies also provide information on the status of SARS-CoV-2 epidemics in situations where record keeping by health systems is not possible^5^.

Infection with SARS-CoV-2 induces diverse humoral and cellular immune responses^6^. Humoral immunity includes antibodies of several immunoglobulin isotypes targeting SARS-CoV-2 proteins, most notably Spike (S) and Nucleocapsid (N). The concentration of antibodies in blood varies substantially between individuals, and with time since infection^6,7,8,9,10,11^. Studies of the duration of immunity to a range of coronaviruses demonstrated that antibodies remain detectable six years after infection, but continue to wane^12^. Longitudinal follow-up of individuals infected with SARS-CoV-2 indicates a pattern of waning of antibody responses consistent with other coronaviruses^13,14^. Within the first three months, antibody levels boost sharply and wane rapidly. Over a longer interval of eight months, antibody levels wane more slowly^6^. These observations can be explained by the bi-phasic nature of antibody kinetics^15^. In the first three months, antibodies are predominantly generated by short-lived plasma cells in secondary lymphoid organs. The long-term response is dominated by antibodies from long-lived plasma cells in the bone marrow. This pattern of bi-phasic waning is observed for infection- and vaccine-induced antibody responses to a wide range of pathogens^16,17,18,19^.

Initial concerns that waning antibody responses would lead to reduced sensitivity of SARS-CoV-2 serological diagnostics over time have unfortunately been confirmed^20^. Using self-administered lateral flow diagnostic tests, a sero-prevalence study of 365,000 adults after the first epidemic wave in the UK observed significantly declining sero-prevalence during July to September 2020^4^. Using a laboratory-based assay for measuring anti-N IgG antibodies, a sero-prevalence survey of blood donors from the Brazilian city of Manaus estimated that 26% of the population had previously been infected^21^. Application of a statistical adjustment to account for antibody waning led to an increased estimate of 76% previously infected. As the pandemic progresses, this problem of declining sensitivity of serological diagnostics is likely to get worse, potentially undermining the utility of sero-prevalence studies.

Serological diagnostics typically classify a sample as positive if a measured antibody level is greater than a defined cutoff. Analysis of quantitative rather than binary antibody levels provides additional information, for example antibody levels are associated with time since infection, symptom severity, and sex^22^. However, large variation in antibody levels between individuals prevents this from having predictive value: detected antibodies could be from a recent infection, or due to immunological memory of an infection that cleared a year ago. This limitation has recently been overcome for a range of pathogens through the combination of multiplex assays and classification algorithms. Using machine learning algorithms to analyse quantitative antibody responses to multiple antigens, the time since previous infection can be estimated for *Plasmodium falciparum* malaria^23,24^, *P. vivax* malaria^25^, and cholera^26^.

In this study, we use multiplex assays to measure antibodies to SARS-CoV-2 in health care workers and hospitalized patients followed for up to eleven months after infection, and apply mathematical models to characterize antibody kinetics in the first year following infection. Classification algorithms were developed that minimize the reduction in the sensitivity of serological tests over time, in addition to estimating time since previous infection from a single blood sample. Finally, we present a method for serological reconstruction of past SARS-CoV-2 transmission using samples from a single cross-sectional survey.

## Methods

### Samples

A panel of 407 negative control serum or plasma samples was assembled from pre-pandemic cohorts (before December, 2019) with ethical approval for broad antibody testing (Table 1). This included 258 serum samples from healthy adult French blood donors, 81 serum samples from Peruvian healthy adult donors, and 69 plasma samples from healthy adult donors from the Thai Red Cross. A panel of 407 positive control serum samples was assembled from individuals with recent SARS-CoV-2 infection. This included 72 samples from patients in Paris hospitals^27,28^, 161 samples from Strasbourg healthcare workers, all confirmed positive by PCR-based testing^29^. Also included were 174 samples from community members of Crépy-en-Valois, France, confirmed seropositive by flow cytometry based testing^30,31^.

**Table 1:** Panels of samples. Age and days post symptom onset are presented as median and ranges.

| cohort | status | participants | samples | PCR positive | age (years) | gender (% male) | days post symptoms |
| --- | --- | --- | --- | --- | --- | --- | --- |
| Établissement Français du Sang 1 | pre-pandemic negative controls | 45 | 45 |  | > 18 |  |  |
| Établissement Français du Sang 2 | pre-pandemic negative controls | 213 | 213 |  | 42 (18,81) | 40% |  |
| Thai Red Cross | pre-pandemic negative controls | 68 | 68 |  | > 18 |  |  |
| Peruvian donors | pre-pandemic negative controls | 81 | 81 |  | > 18 |  |  |
| Hôpital Bichat (Paris, France) | hospitalized patients | 2 | 8 | 2 | 31 (30, 32) | 100% | 14 (8, 24) |
| Hôpital Cochin (Paris, France) | hospitalized patients | 64 | 64 | 64 | 55 (25, 79) | 76% | 17 (10, 28) |
| Strasbourg hospitals 1 | infected healthcare workers | 161 | 161 | 161 | 32 (20, 62) | 31% | 24 (13, 39) |
| Crépy-en-Valois community | infected community members (flow cytometry positive) | 154 | 174 | 0 | 17 (15, 56) | 34% |  |
| Dublin hospitals | hospitalized patients | 194 | 213 | 194 | 55 (21, 92) | 47% | 13 (1, 126) |
| Strasbourg hospitals 2 | follow-up of infected healthcare workers | 347 | 724 | 347 | 41 (21,74) | 23% | 132 (11, 284) |
| Institut Mutualiste Montsouris (Paris, France) | sero-prevalence survey in healthcare workers (unknown status) | 769 | 769 | 20 | 41 (18, 72) | 27% |  |
| Institut Mutualiste Montsouris (Paris, France) | follow-up of sero-positive healthcare workers | 29 | 29 | 12 | 37 (24, 63) | 41% | 304 (285, 336) |

The duration of antibody responses following SARS-CoV-2 infection was studied in longitudinal cohorts of hospitalized patients and healthcare workers. 213 serum samples from 194 patients in Dublin hospitals were collected, with date post symptom onset extending to four months. 724 serum samples from 347 healthcare workers in Strasbourg hospitals were collected, with date post symptom onset extending to nine months^32^.

In April 2020, our team implemented a study of the sero-prevalence of SARS-CoV-2 in healthcare workers from Institut Mutualiste Montsouris, a hospital in Paris. Serum samples were collected from 769 healthcare workers, and tested with our multiplex assay (Appendix Figure 1). Healthcare workers who tested sero-positive in April 2020 were invited to present a second sample in January 2021. In total we obtained follow-up samples from 29 healthcare workers.

### Serological assays

We set up a 9-plex bead-based assay allowing simultaneous detection of antibody responses to five SARS-CoV-2 antigens and four seasonal coronaviruses (Spike proteins of NL63, 229E, HKU1, OC43) in 1µL serum or plasma samples that were heat-inactivated at 56°C for 30 minutes. SARS-CoV-2 antigens were from Spike (whole trimeric Spike (S), its Receptor Binding Domain (RBD) and Spike subunit 2 (S2)), nucleocapsid protein (N) and a Membrane-Envelope fusion protein (ME). ME and S2 antigens were purchased from Native Antigen, Oxford, UK and all other antigens were produced as recombinant proteins at Institut Pasteur. The mass of proteins coupled on beads was optimized to generate a log-linear standard curve with a pool of 27 positive sera prepared from RT-qPCR-confirmed SARS-CoV-2 patients. We measured the levels of IgG, IgA and IgM of each sample in three separate assays. Briefly, serum was incubated with mixed antigen-coupled beads for 30 minutes at a 1/200 final dilution for IgG or 1/400 for IgA and IgM. Secondary antibodies conjugated to R-phycoerythrin (Jackson Immunoresearch) were used at 1/120, 1/200 or 1/400 for detection of specific IgG, IgA and IgM respectively. All dilutions and cycles of washing steps were done in phosphate buffer saline supplemented with 1% bovine serum albumin and 0.05% (v/v) Tween-20. On each assay plate, two blanks (only beads, no serum) were included to control for background signal as well as a standard curve prepared from two-fold serial dilutions (1/50 to 1/102,400) of a pool of positive controls. Plates were read using a Luminex® MAGPIX® system and the median fluorescence intensity (MFI) was used for analysis. A 5-parameter logistic curve was used to convert MFI to relative antibody unit (RAU), relative to the standard curve performed on the same plate to account for inter-assay variations.

The data from our multiplex assay was compared against data from two different neutralization assays with live virus using a subset of serum samples. Firstly, we implemented an S-Fuse assay as described elsewhere^32^. Neutralization activity was measured as the reciprocal dilution required to obtain a 50% reduction in neutralization (IC50). Secondly, we implemented a foci reduction neutralization test (FRNT) based on the detection of neutralizing antibodies directed against SARS-CoV-2. This assay was performed under BSL-3 conditions as it facilitates infection of African green monkey kidney cells (VeroE6; ATCC CRL-1586) with live-virus of a Cambodian SARS-CoV-2 isolate (GISAID: EPI_ISL_411902). Infection is visualized 14-16h after inoculation by staining of infected cells with a SARS-CoV-2 specific antibody (# ABIN1030641), targeting the S2 subunit of the viral spike protein. Neutralizing antibody titers are expressed as the reciprocal serum/plasma dilution that induces 50% reduction of infection (FRNT50) and is calculated by log probit regression analysis.

### Mathematical model of antibody kinetics

SARS-CoV-2 antibody kinetics are described using a previously published mathematical model of the immunological processes underlying the generation and waning of antibody responses following infection^33^.

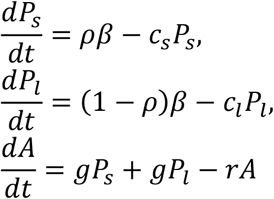

β denotes the boost in antibody secreting plasma cells. It is assumed that a proportion *ρ* of plasma cells are short-lived (*P*_*s*_) waning at rate *c*_*s*_, and a proportion 1 – *ρ* are long-lived (*P*_*l*_) waning at rate *c*_*l*_. Plasma cells generate antibodies (IgG, IgM or IgA) at rate *g*, and *r* is the rate of decay of antibody molecules.

Statistical inference was implemented within a mixed-effects framework allowing for characterisation of the kinetics within all individuals while also describing the population-level patterns (Statistical Appendix). On the population level, both the mean and variation in antibody kinetics are accounted for. Models were fitted in a Bayesian framework using Markov chain Monte Carlo methods with priors informed by estimates from long-term studies of antibody kinetics following infection with other coronaviruses^20^.

### Classification algorithms

Measurements of antibodies of three isotypes (IgG, IgM, IgA) to multiple SARS-CoV-2 antigens were used to create a training dataset. Samples where the time post symptom onset was ≤ 14 days or unknown were excluded. In total we had 407 samples from pre-pandemic negative controls and 1402 positive samples. In a first step, a Random Forests binary classification algorithm was developed. A threshold corresponding to 99% specificity was selected. In a second step, a Random Forests multiway classification algorithm was developed for categorizing samples into four classes: (i) negative; (ii) infected ≤ 3 months ago; (iii) infected 3 – 6 months ago; and (iv) infected 6 – 12 months ago. The algorithm was calibrated to have 99% specificity for correctly classifying negative samples. Positive samples were then classified according to the maximum number of votes. Uncertainty in classification performance was assessed via 1000-fold cross-validation with a training set comprising two thirds of the data and a disjoint testing set comprising a third of the data. Classification algorithms were implemented in R (version 3.4.3).

### Statistical methods for serological surveillance

Imperfect diagnostic sensitivity causes a downwards bias in sero-prevalence estimates, whereas imperfect specificity causes an upwards bias in seroprevalence estimates. This bias can be corrected for using a Rogan-Gladen estimator, with incorporation of cross-validated uncertainty^34^. Multiway classification algorithms provide estimates of the proportion of a population infected during different time intervals. These estimates are subject to biases which can be adjusted for using a multivariate extension of the Rogan-Gladen estimator described in the Statistical Appendix.

### Ethics

Serum samples were biobanked at the Clinical Investigation and Access to BioResources platform at Institut Pasteur (Paris, France). Samples were obtained from consenting individuals through the CORSER study (NCT04325646), directed by Institut Pasteur and approved by the Comité de Protection des Personnes Ile de France III, and the French COVID cohort (NCT04262921), sponsored by Inserm and approved by the Comité de Protection des Personnes Ile de France VI. Samples from French blood donors were approved for use by Etablissement Français du Sang (Lille, France) and approved through the CORSER study by the Comité de Protection des Personnes Ile de France VI. Sample collection in Hôpital Cochin was approved by the Research Ethics Commission of Necker-Cochin Hospital. Samples from healthcare workers in Strasbourg University Hospitals followed longitudinally were collected as part of an ongoing clinical trial (ClinicalTrials.gov Identifier: NCT04441684) which received ethical approval from the Comité de Protection des Personnes Ile de France III. Samples collected from patients in Dublin received ethical approval for study from the Tallaght University Hospital (TUH)/St James’s Hospital (SJH) Joint Research Ethics Committee (reference REC 2020–03). Use of the Peruvian negative controls was approved by the Institutional Ethics Committee from the Universidad Peruana Cayetano Heredia (SIDISI 100873). The Human Research Ethics Committee at the Walter and Eliza Hall Institute of Medical Research and the Ethics Committee of the Faculty of Tropical Medicine, Mahidol University, Thailand, approved the use of the Thai negative control samples. Informed written consent was obtained from all participants or their next of kin in accordance with the Declaration of Helsinki.

## Results

### SARS-CoV-2 antibodies over time

Antibody responses of three isotypes (IgG, IgM, IgA) to nine coronavirus antigens were measured in 407 pre-pandemic serum samples, and 1402 serum samples from individuals with previous SARS-CoV-2 infection. 961/1402 of the positive samples were from individuals with SARS-CoV-2 infection confirmed by PCR-based testing with available data on time post symptoms. Figure 1 presents the IgG, IgM and IgA antibody responses to SARS-CoV-2 S, RBD, and N measured over time. Appendix Figures 2 and 3 presents the antibody responses to S2, ME, and the Spike proteins of the four human seasonal coronaviruses. Notably there is substantial inter-individual variation in antibody responses, with antibody levels varying by orders of magnitude between individuals. As a measure of functional immunity, we applied live virus neutralizing assays to a subset of samples. Substantial variation in neutralizing activity between individuals and over time (Appendix Figure 4).

**Figure 1:**
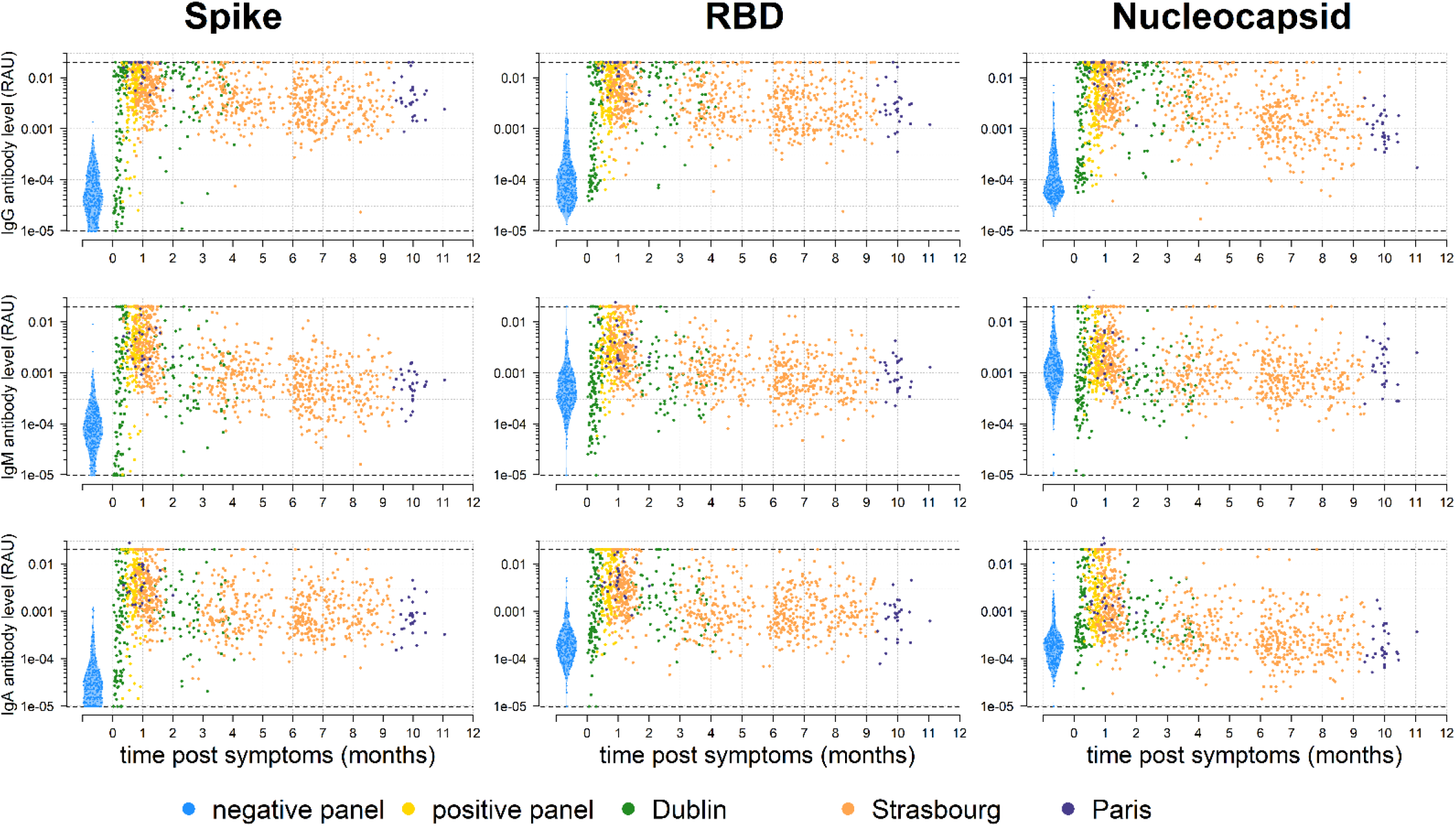
Antibody kinetics in the first year following infection with SARS-CoV-2. A bead-based multiplex Luminex assay used to measure antibodies of multiple isotypes (IgG, IgM, IgA) to multiple antigens in serum samples from individuals with PCR-positive SARS-CoV-2 infection and pre-pandemic negative controls.

### Modelled SARS-CoV-2 antibody kinetics

Our data on measured SARS-CoV-2 antibody responses was supplemented with data from six other longitudinal studies of the SARS-CoV-2 antibody response, and one longitudinal study of the SARS-CoV-1 antibody response. Appendix Figure 5 provides a comparison of the measured antibody responses between the eight studies. The data have been rescaled so that the average antibody response for each study equals one at day 14 after symptom onset. In addition to the large inter-individual variation, there is notable variation in antibody levels between studies. For example, the large dynamic range in the study by Iyer *et al*^8^, reveals the boosting and subsequent decay of anti-S IgG antibodies in the first two months post infection, a phenomenon that is missed by assays without a comparably high upper limit of detection.

A mathematical model of antibody kinetics was simultaneously fitted to data from all eight studies. Appendix Figure 6 provides an overview of the fit of the model to the data. Figure 2a provides example fits to data from one individual from each of the eight studies. Figure 2b provides model predictions of the percentage antibody level remaining over the first two years post infection, where 100% is assumed to be the antibody response at day 14 following symptom onset. There are considerable differences in the pattern of waning between isotypes and between antigens. Table 2 summarises the duration of the antibody response. One year following symptom onset, we estimate that 36% (95% range: 11%, 94%) anti-S IgG remains, 31% (9%, 89%) anti-RBD IgG remains, and 7% (1%, 31%) anti-N IgG remains. The uncertainty represents the considerable degree of inter-individual variation in the duration of the antibody responses. Antibodies of the IgM isotype waned more rapidly, with 6% (0%, 27%) anti-S IgM remaining after one year, 9% (2%, 32%) anti-RBD IgM remaining after one year, and 15% (4%, 50%) anti-N IgM remaining after one year. Antibodies of the IgA isotype also waned rapidly, with 18% (4%, 67%) anti-S IgA remaining after one year, 10% (3%, 38%) anti-RBD IgA remaining after one year, and 3% (0%, 13%) anti-N IgA remaining after one year. We also observed comparable reductions in titres for viral neutralization over time (Appendix Figure 4), although the small number of samples prevented application of our model.

**Figure 2:**
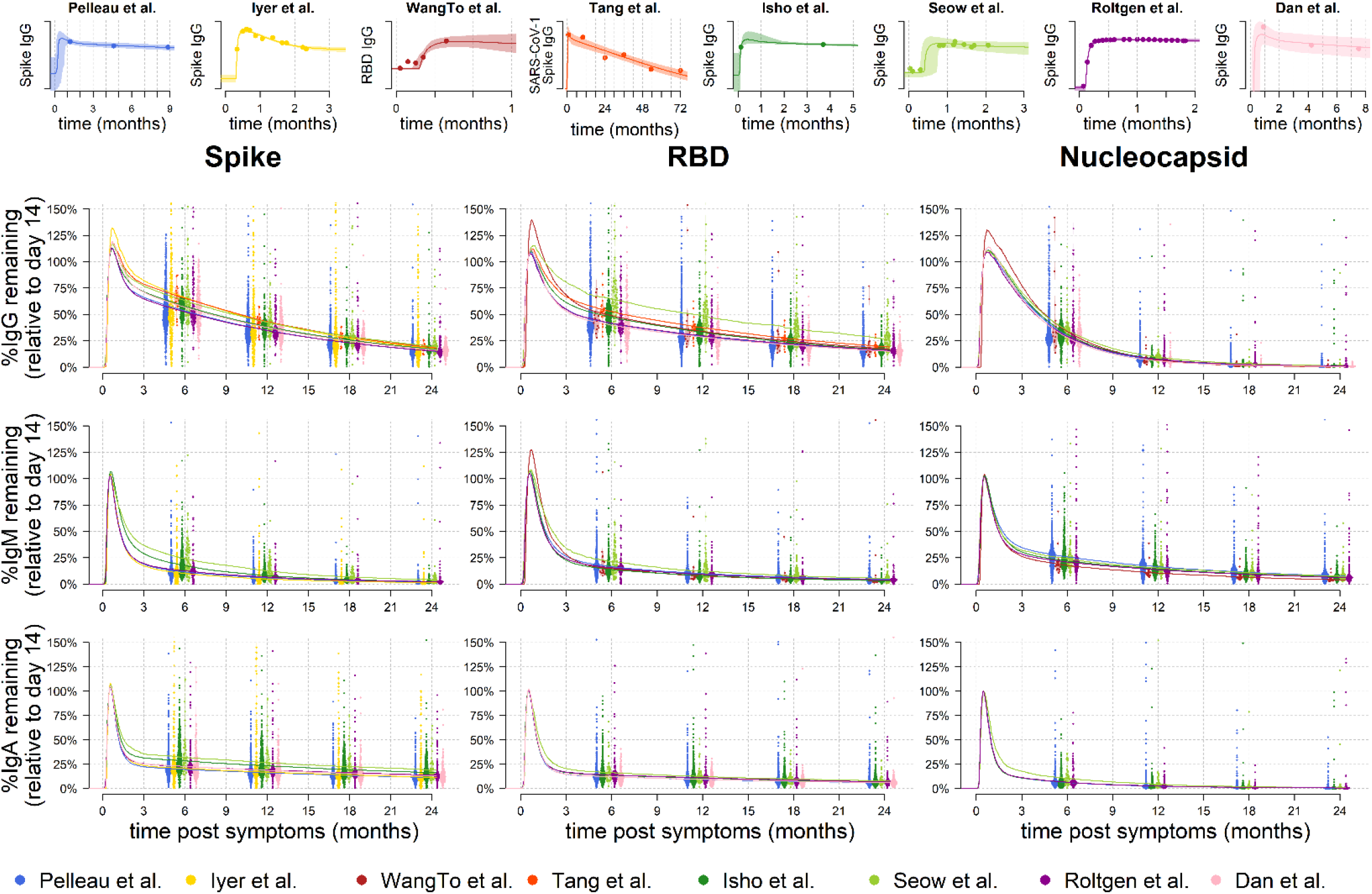
Modelled SARS-CoV-2 antibody kinetics. A mathematical model of SARS-CoV-2 antibody kinetics was simultaneously fitted to data from seven studies of SARS-CoV-2 and one study of SARS-CoV-1. **(a – top row)** Examples of the model fit to the data for one individual from each study. Data are represented as points, posterior median model prediction as lines, and 95% credible intervals as shaded areas. **(b – middle and bottom rows)** Model predicted duration of antibodies within the first 2 years following infection. Antibody levels are shown relative to the expected antibody level at day 14 following symptom onset. Each point represents the prediction from an individual at 6, 12, 18 and 24 months post symptoms. The median predictions for each of the eight studies are presented as lines.

**Table 2:**
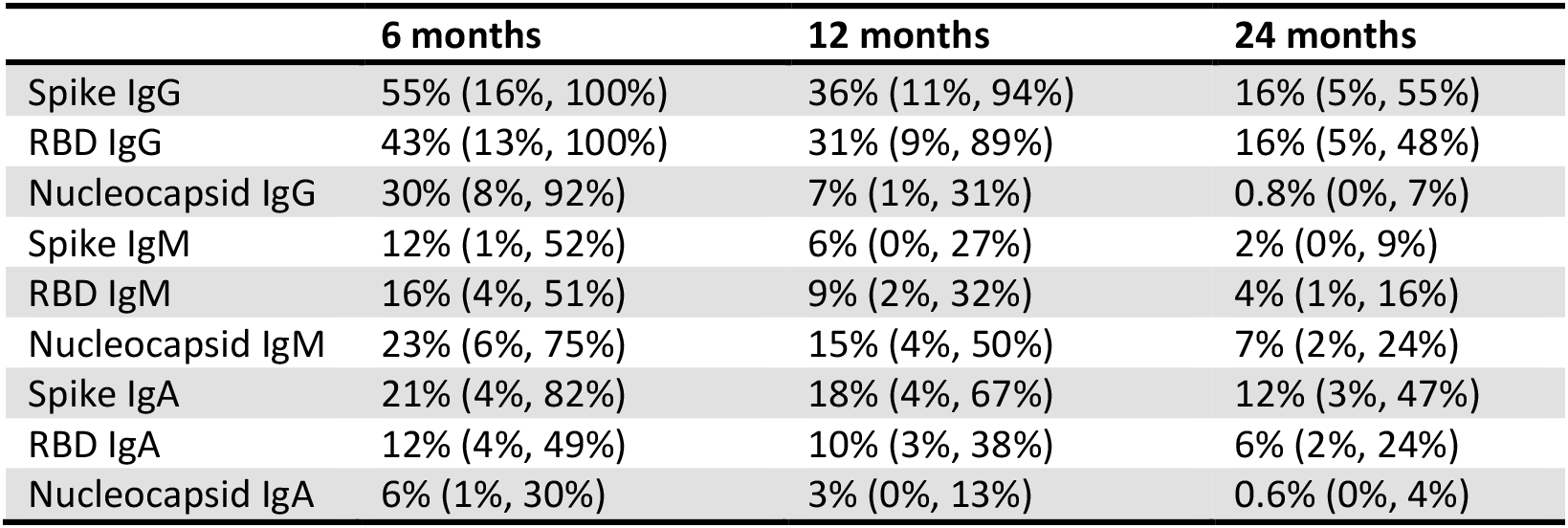
Estimated duration of antibody responses following SARS-CoV-2 infection. The percentage antibody level remaining over time is compared to the measured antibody level 14 days after symptom onset. Estimates are presented as the population median, with the 95% range due to inter-individual variation.

|  | 6 months | 12 months | 24 months |
| --- | --- | --- | --- |
| Spike IgG | 55% (16%, 100%) | 36% (11%, 94%) | 16% (5%, 55%) |
| RBD IgG | 43% (13%, 100%) | 31% (9%, 89%) | 16% (5%, 48%) |
| Nucleocapsid IgG | 30% (8%, 92%) | 7% (1%, 31%) | 0.8% (0%, 7%) |
| Spike IgM | 12% (1%, 52%) | 6% (0%, 27%) | 2% (0%, 9%) |
| RBD IgM | 16% (4%, 51%) | 9% (2%, 32%) | 4% (1%, 16%) |
| Nucleocapsid IgM | 23% (6%, 75%) | 15% (4%, 50%) | 7% (2%, 24%) |
| Spike IgA | 21% (4%, 82%) | 18% (4%, 67%) | 12% (3%, 47%) |
| RBD IgA | 12% (4%, 49%) | 10% (3%, 38%) | 6% (2%, 24%) |
| Nucleocapsid IgA | 6% (1%, 30%) | 3% (0%, 13%) | 0.6% (0%, 4%) |

### Estimating time since previous SARS-CoV-2 infection

Using a dataset on measured IgG, IgM and IgA antibody levels from pre-pandemic negative controls and samples from individuals with SARS-CoV-2 infection confirmed by PCR-based testing and followed for up to eleven months after symptom onset, Random Forests binary classification algorithms were trained to identify individuals with previous SARS-CoV-2 infection (Appendix Figures 7 and 8). Next, a Random Forests multi-way classification was trained to simultaneously identify previous infection and estimate the time since infection (Figure 3). The diagnostic test identified samples from individuals with previous SARS-CoV-2 infection with 99% specificity and 98% (95% confidence interval: 94%, 99%) sensitivity (Figure 3a). The diagnostic test classified samples from individuals infected within the previous 3 months (Figure 3b). Notably, it was easier to distinguish recent infections (<3 months) from older infections (6 – 12 months), compared to infections that occurred 3 – 6 months ago. There was limited statistical signal to distinguish between infections that occurred 3 – 6 months ago, and older infections occurring more than 6 months ago (Figure 3c). A breakdown of classification performance by time since infection is provided in Figure 3D. The diagnostic test accurately classifies samples of all categories, with the exception of samples from individuals infected 3 – 6 months ago. Many of these samples were incorrectly classified in the neighbouring infection categories of 0 – 3 months or 6 – 12 months.

**Figure 3:**
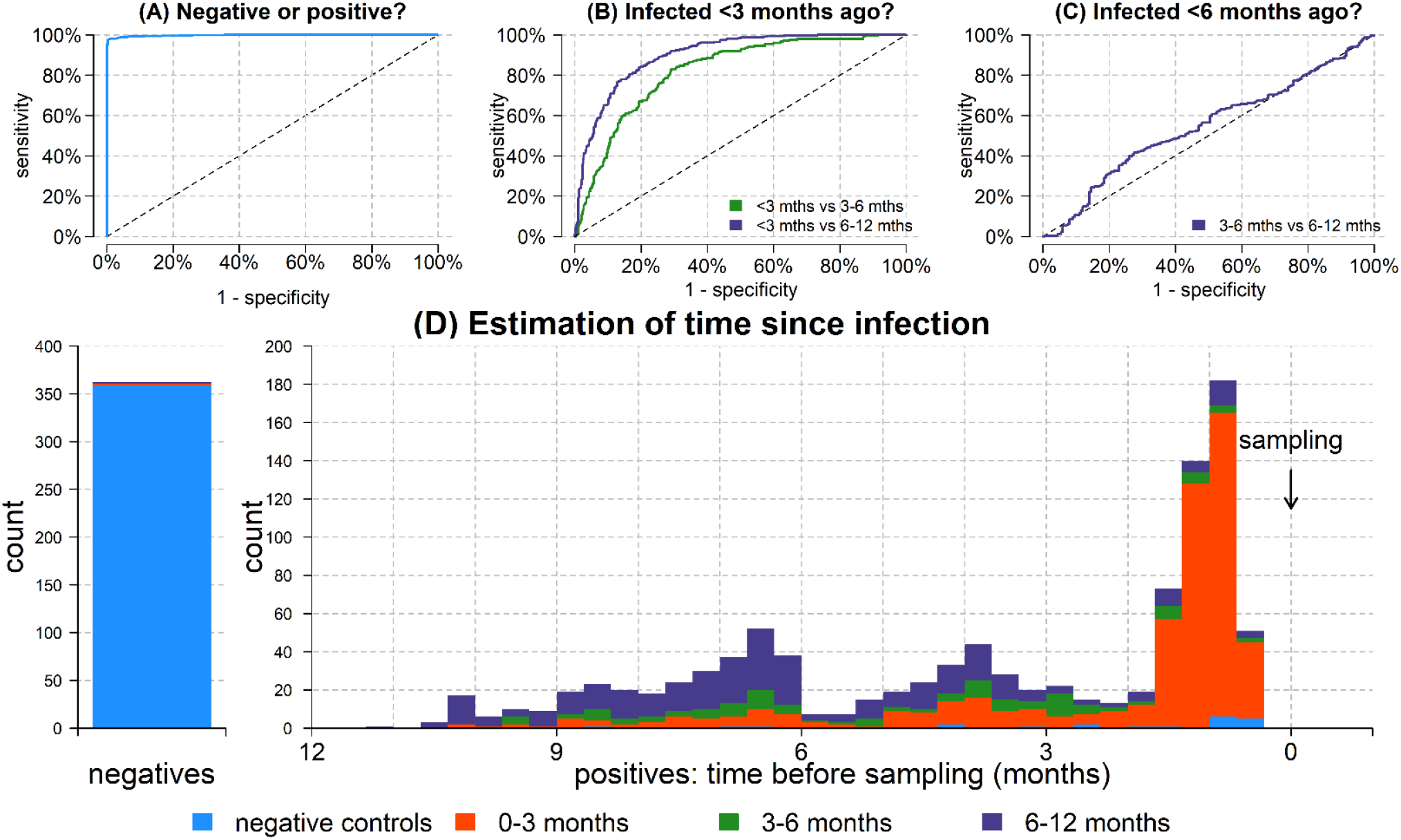
Classification of time since previous SARS-CoV-2 infection. A cross-validated multi-way classification algorithm was trained to estimate time since infection. **(A)** The algorithm can differentiate between positive and negative samples. **(B)** The algorithm can classify individuals infected within the previous 3 months. **(C)** There is limited diagnostic power to distinguish between infections that occurred 3-6 months ago versus 6-12 months ago. **(D)** Breakdown of classification performance according to time since previous infection. Colours represent model predicted classification. >99% of negative samples are correctly classified as negative (blue). For the positive samples, the distribution shows the time since previous infection. Samples with time since infection <3 months are mostly classified in the 0 – 3 month category (red). Samples with time since infection >6 months ago are mostly classified in the 6 – 12 month category (purple). There is a substantial degree of misclassification of samples with time since infection 3 – 6 months ago. This is due to the temporal imbalance in the training data.

### Serological reconstruction of COVID-19 epidemics

When applied to samples from a cross-sectional sero-prevalence survey, a diagnostic test capable of estimating time since infection can provide a serological reconstruction of past SARS-CoV-2 epidemics. Figure 4 presents four simulated epidemics where 1,000 individuals were sampled, and 40% were previously infected. A range of scenarios were simulated, including a recent epidemic wave, an older epidemic wave, a double wave epidemic, and constant transmission. The algorithm was able to accurately reconstruct all epidemic profiles. For example, in the recent wave scenario 60% of the population were negative and the model estimated 59.8% (95% CI: 59.3%, 61.4%); 35.8% were infected 0 – 3 months ago and the model estimated 35.5% (95% CI: 25.7%, 40.3%); 4.2% were infected 3 – 6 months ago and the model estimated 4.2 % (95% CI: 0.0%, 14.4%); and 0% were infected 6 – 12 months ago and the model estimated 0% (95% CI: 0.0%, 3.3%).

**Figure 4:**
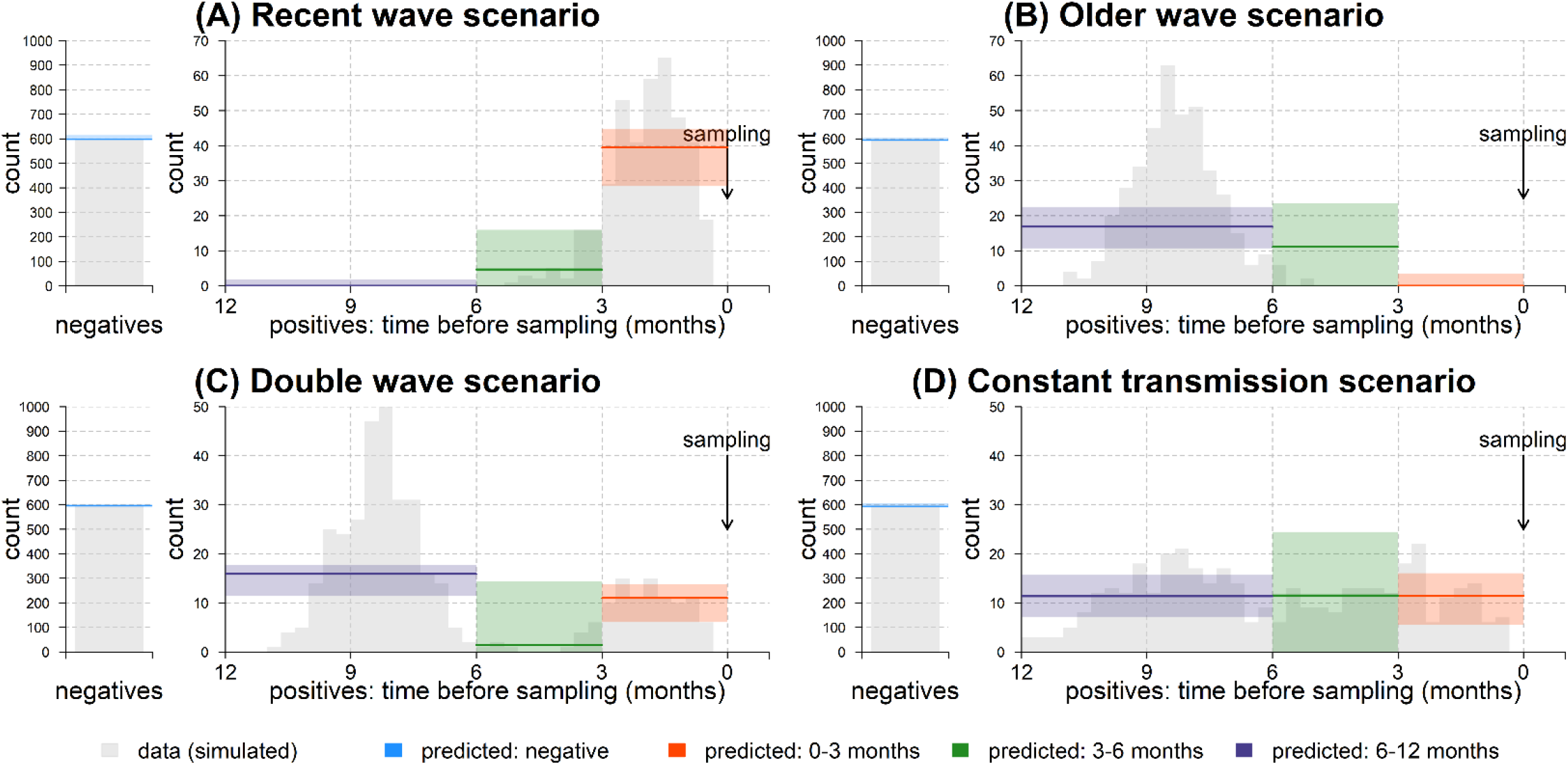
Serological reconstruction of previous COVID-19 epidemics. The grey histograms represent simulated data from a range of SARS-CoV-2 epidemic scenarios. Data were simulated from 1000 samples from a cross-sectional sero-prevalence study. It was assumed that 600 samples were negative, with 400 positive samples with varying time since infection. For previously infected individuals, the histograms represent the time since infection. Based on antibody levels sampled at a single time point, our computational method was used to provide a serological reconstruction of previous COVID-19 epidemics. The solid lines represent the model predictions for the categories of negative; infected 0 – 3 months ago; infected 3 – 6 months ago; and infected 6 – 12 months ago. Shaded regions represent 95% confidence intervals.

## Discussion

Infection with SARS-CoV-2 induces a complex, diverse immune response, that varies by orders of magnitude between individuals, and changes over time. This diversity is a challenge to immunologists and vaccinologists, but presents an opportunity to diagnostic developers armed with multiplex assays and machine learning algorithms. By quantifying the kinetics of different components of the humoral immune response, it is possible to provide classification of previous infection that minimizes the reduction of diagnostic sensitivity over time, and also allows estimation of the time since infection.

Based on data from a range of studies with up to eleven months follow up after symptom onset, we estimate that 31% (95% CrI: 9%, 89%) of anti-RBD IgG antibody levels remain one year after infection. Antibodies of other isotypes waned more rapidly, with 9% (95% CrI: 2%, 32%) of anti-RBD IgM antibody levels remaining after one year, and 10% (95% CrI: 3%, 38%) of anti-RBD IgA antibody levels remaining after one year. There was considerable variation in kinetic profiles between different SARS-CoV-2 antigens, with 7% (95% CrI: 1%, 31%) of anti-N IgG antibody levels remaining after one year. Although the determinants of the duration of antigen-specific antibody responses remain poorly understood^22^, the diversity in patterns of antibody kinetics can be quantified using epidemiological studies, yielding valuable information for serological diagnostics.

The majority of commercially available serological tests classify individuals as having previous SARS-CoV-2 infection if a measured antibody response is greater than a defined cutoff^35^. Instead of reducing a complex antibody response to a binary data point, a mode detailed serological signature based on quantitative measurements of multiple antibody responses provides two notable advantages^20^. Firstly, the reduction in diagnostic sensitivity associated with waning antibodies is minimized – no reduction in diagnostic sensitivity over the eleven months of follow up was observed. Secondly, the time since previous infection can be estimated providing valuable additional epidemiological information. For the current assay, previous infections were categorised into intervals of 0 – 3 months, 3 – 6 months, and 6 – 12 months. More precise classification is possible in theory, but this must be balanced against the statistical signal. For example, there was little statistical signal to discriminate between infections that occurred 6 – 9 months ago, versus infections that occurred 9 – 12 months ago. It is anticipated that further improvements to the assay such as the incorporation of new antigens, more training samples with a range of time since infection, and better algorithms will lead to improvements in accuracy.

Existing sero-prevalence studies estimate the proportion of a population previously infected with SARS-CoV-2. The addition of a diagnostic tool capable of estimating time since infection allows for the serological reconstruction of past transmission trends. Thus, using samples from a sero-prevalence study collected at a single time point, we can discriminate between a scenario of constant SARS-CoV-2 transmission and a scenario where transmission occurs in distinct epidemic waves. This diagnostic technology has a range of possible applications. For countries that experienced double wave SARS-CoV-2 epidemics, it has been challenging to quantify the relative magnitude of these waves due to the time required to scale up PCR-based diagnostic testing^36^. Furthermore, there are many epidemiological settings where it is unknown if SARS-CoV-2 transmission was constant over time, or occurred as distinct epidemic waves^5^.

The widespread introduction of SARS-CoV-2 vaccines will lead to vaccine-induced immunity in many individuals. The majority of vaccines against SARS-CoV-2 target the Spike protein’s RBD. As our diagnostic assay and algorithms include Spike and RBD antigens, serum samples from vaccinated individuals would likely be classified as positive. However, by measuring antibody responses to Nucleocapsid, Membrane, Envelope, and other viral proteins^37^ it will be possible to modify the diagnostic test to provide three-way classification: negative; previously infected; or vaccinated. Identification of individuals who have been both infected and vaccinated will be challenging. Having classified an individual as previously infected or vaccinated, the diagnostic algorithm can then be used to estimate time since infection or time since vaccination. Such an approach could contribute to efforts to measure population-level immunity to SARS-CoV-2, whether induced by infection or vaccination.

There are several limitations to this study. Estimates of the duration of antibody responses are based on data from multiple studies, each using a unique immunoassay^6,7,8,9,10,11,12^. Every immunoassay may differ in terms of background reactivity, cross-reactivity with other pathogens, protein formulation, dynamic range and reproducibility. We believe that the benefit of drawing on multiple data sources outweighs the benefit of having a smaller more homogeneous database, especially since the mathematical model of antibody kinetics is sufficiently flexible to incorporate data from multiple assays. Our selection of a mechanistic mathematical model of antibody kinetics is a potential limitation. The model is based on a mechanistic understanding of the immunological processes underlying the generation and persistence of antibodies, and imposes a flexible functional form on how antibody levels change over time. Although this approach has been validated in a range of applications^16,17,18,19^, there will be instances where the model fails to capture the true pattern of antibody kinetics, for example in immune-deficient individuals. An advantage of a mechanistic model versus a non-parametric statistical model is the ability to make projections forward in time. We have provided predictions up to two years following infection, for example by estimating that 16% (5%, 48%) of anti-RBD IgG antibodies remain after two years. There is a risk to providing predictions beyond the timescale of the data, but these predictions can be easily falsified via continued longitudinal studies.

The diagnostic assay is not exempt from the challenges of antibody waning. Although no reductions in diagnostic sensitivity over time were observed, reduced sensitivity will likely be observed as we analyse additional samples over longer durations of follow up. There are substantial challenges in providing estimates of time since infection. Although the approach can reliably reconstruct the distribution of infection times across a population, there will be substantial uncertainty in estimates of the time since previous infection for individual samples.

Sero-prevalence studies are playing a critical role in monitoring the progress of the SARS-CoV-2 pandemic. In the early stages of the pandemic, immunoassays had the advantage of measuring high antibody levels in the initial months after infection. As the pandemic progresses, sero-prevalence will become more challenging to accurately measure due to waning antibody responses and increased vaccine-induced immunity. Multiplex assays and algorithms accounting for how antibody levels change over time may be an important tool for ensuring the ongoing utility of sero-surveillance.

## Data Availability

Data and code will be made available online following peer review of the article. In the meantime, requests for data or code should be made to the corresponding author.

## Acknowledgements

The French COVID cohort is supported by the REACTing consortium and by the French Directorate General for Health. Jessica Vanhomwegen (Institut Pasteur) is thanked for provision of a MAGPIX machine. Jérôme Hadjadj and Laura Barnabei (Institut Imagine, Paris) are thanked for their work on the Hôpital Cochin study. Dionicia Gamboa (Cayetano Heredia University, Lima) is thanked for sharing negative control samples from Peru. Jetsumon Sattabongkot (Mahidol University, Bangkok) is thanked for sharing negative control samples from Thailand. Marie-Noelle Ungeheuer and Blanca Liliana Perlaza are thanked for processing samples at the Clinical Investigation and Access to BioResources platform in Institut Pasteur. We thank all patients and health-care workers who kindly agreed for samples to be used for medical research purposes.

## Appendix Figures

**Appendix Figure 1:**
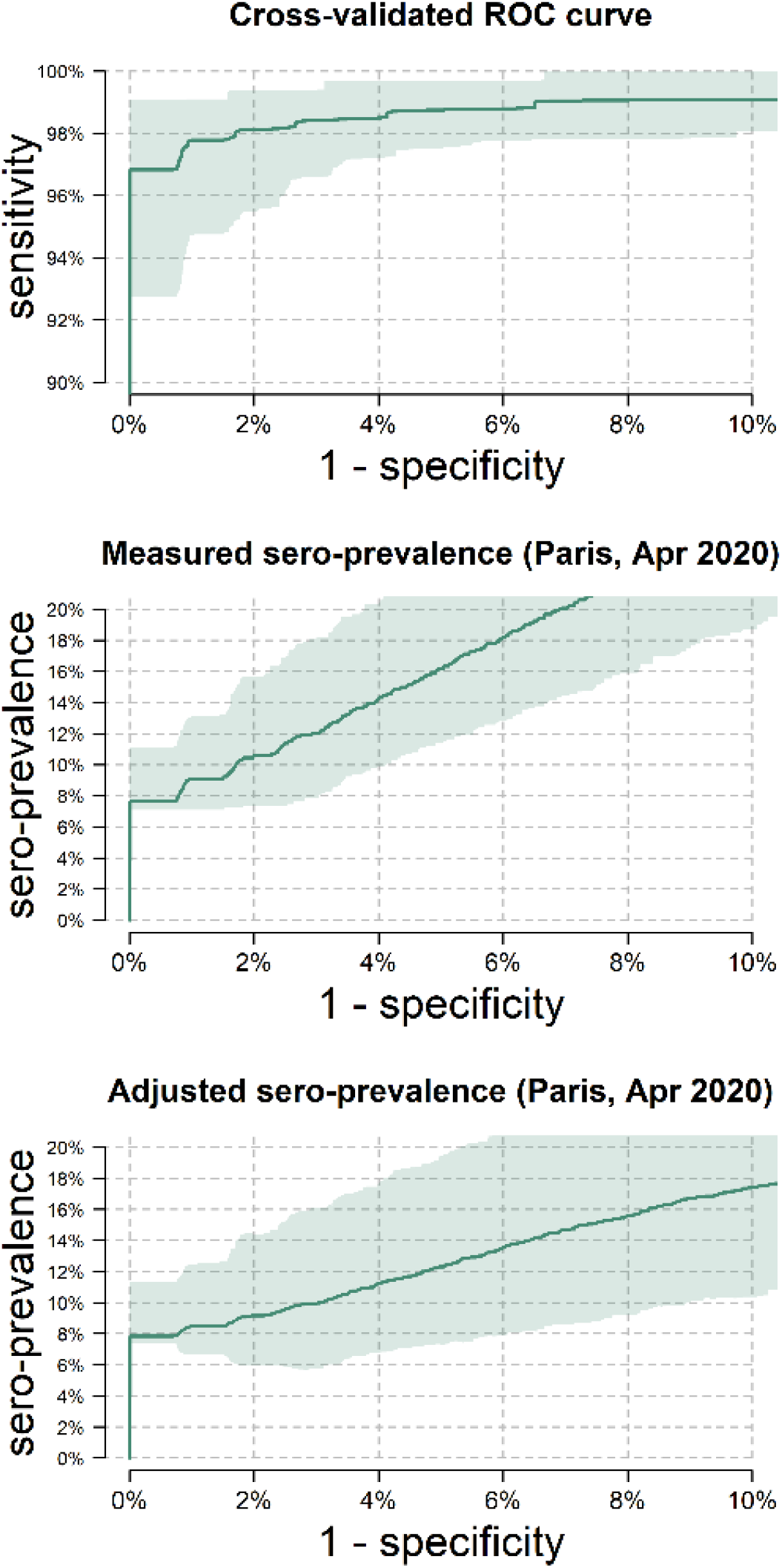
Application of multiplex assay to a sero-prevalence study in Institut Mutualiste Montsouris hospital in Paris. **(A)** Cross-validated receiver operating characteristic (ROC) curve showing the diagnostic performance of a multiplex assay for measuring IgG, IgM and IgA antibodies to SARS-CoV-2 antigens. This assay can achieve 98% sensitivity and 99% specificity. **(B)** Measured sero-prevalence in 769 healthcare workers across a range of diagnostic specificities. **(C)** Sero-prevalence adjusted with a Rogan-Gladen estimator. It can be shown that accuracy is maximized by selecting high specificity^20^. At 99% specificity, this gives an estimated sero-prevalence of 8.7% (95 CI: 6.7%, 11.0%). In all panels, the green line represents the median prediction and the shaded regions represent the 95% confidence intervals.

**Appendix Figure 2:**
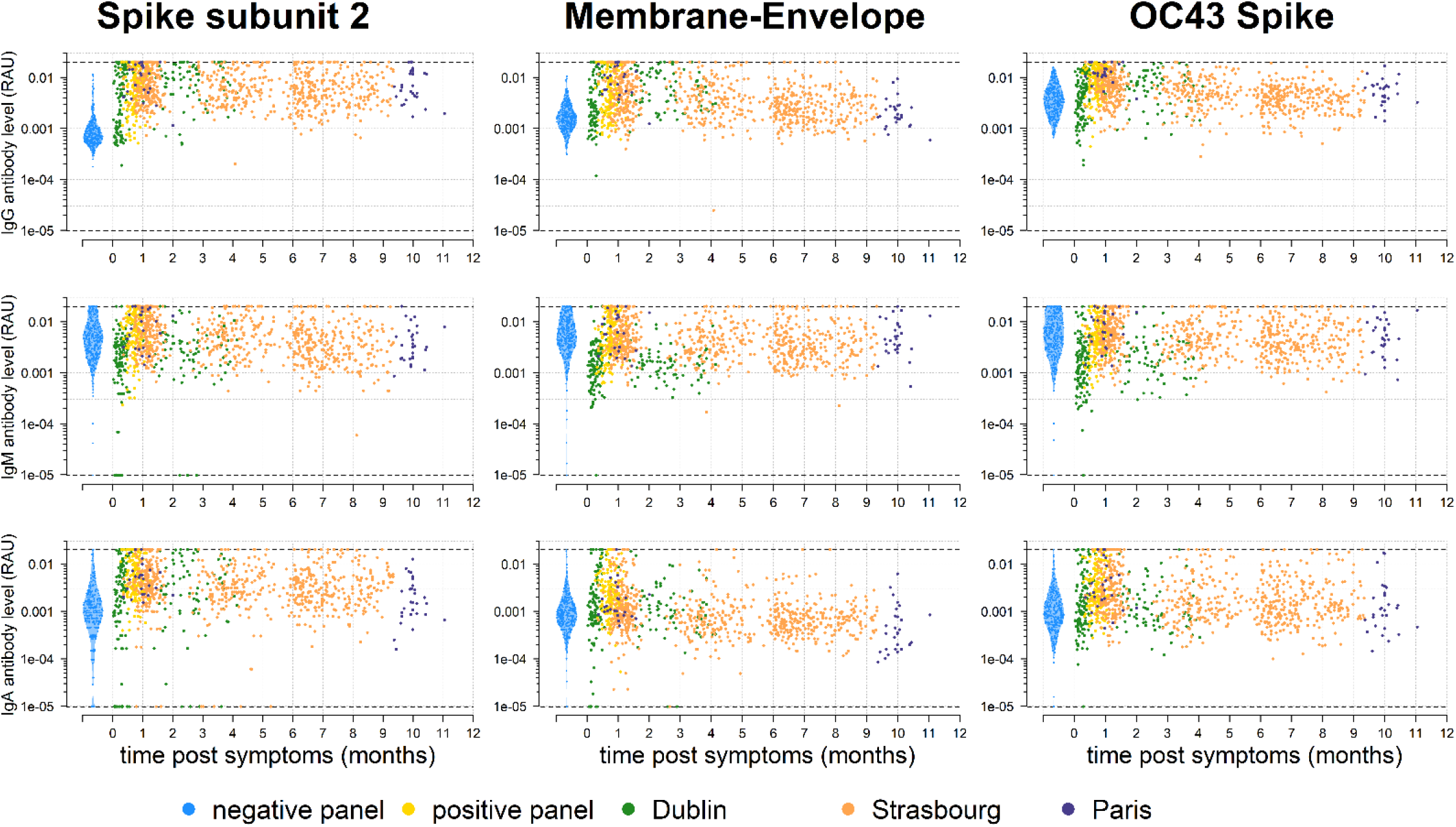
Antibody kinetics in the first year following infection with SARS-CoV-2. A bead-based multiplex Luminex assay was used to measure antibodies of multiple isotypes (IgG, IgM, IgA) to multiple antigens in serum samples from individuals with PCR-positive SARS-CoV-2 infection and pre-pandemic negative controls. Shown here are the antibody responses to the SARS-CoV-2 Spike subunit 2 (S2) and Membrane-Envelope fusion (ME) antigens, and the Spike protein of the OC43 human seasonal coronavirus.

**Appendix Figure 3:**
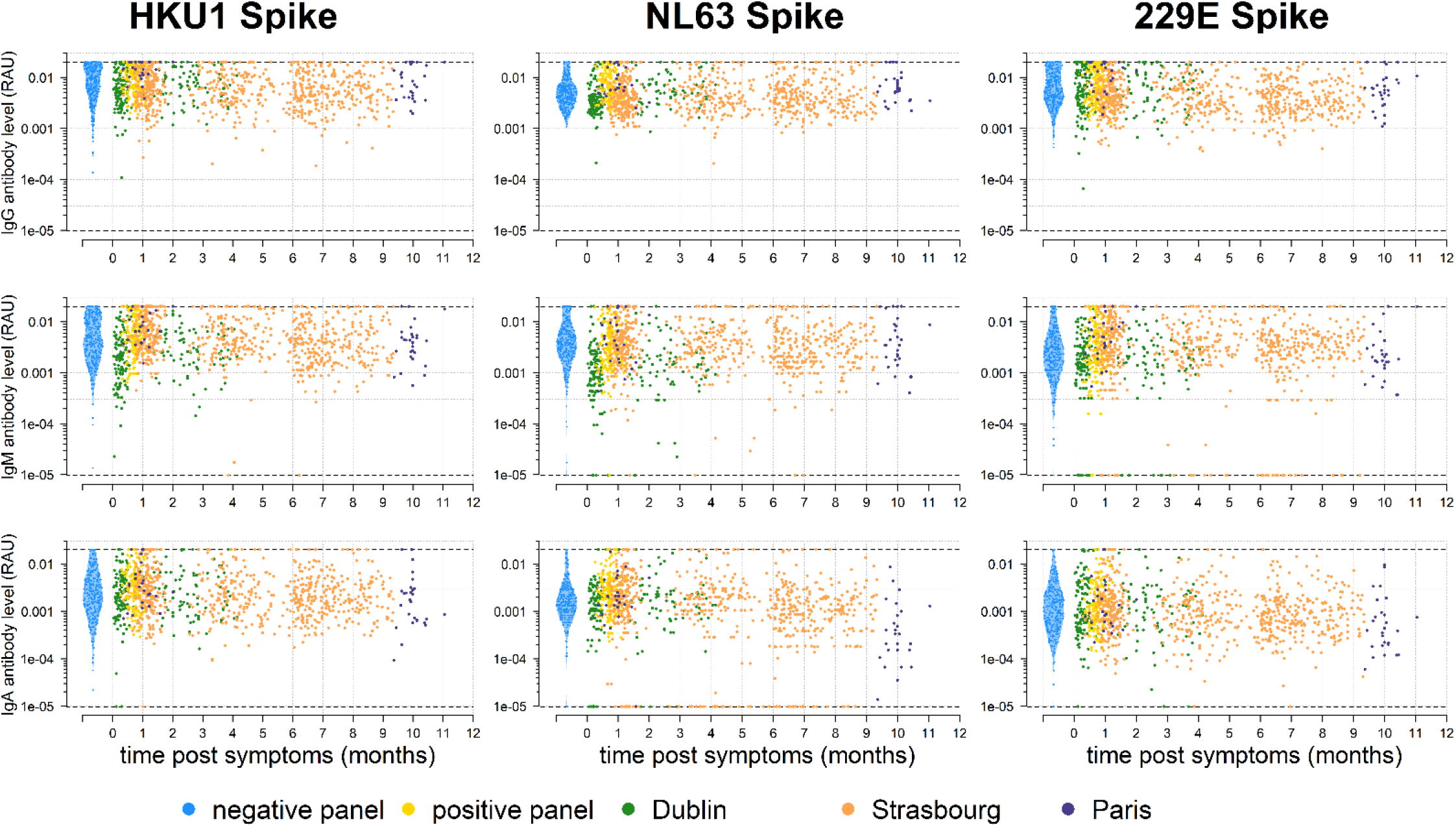
Antibody kinetics in the first year following infection with SARS-CoV-2. A bead-based multiplex Luminex assay was used to measure antibodies of multiple isotypes (IgG, IgM, IgA) to multiple antigens in serum samples from individuals with PCR-positive SARS-CoV-2 infection and pre-pandemic negative controls. Shown here are the antibody responses to the Spike proteins of the HKU1, NL63 and 229E human seasonal coronaviruses.

**Appendix Figure 4:**
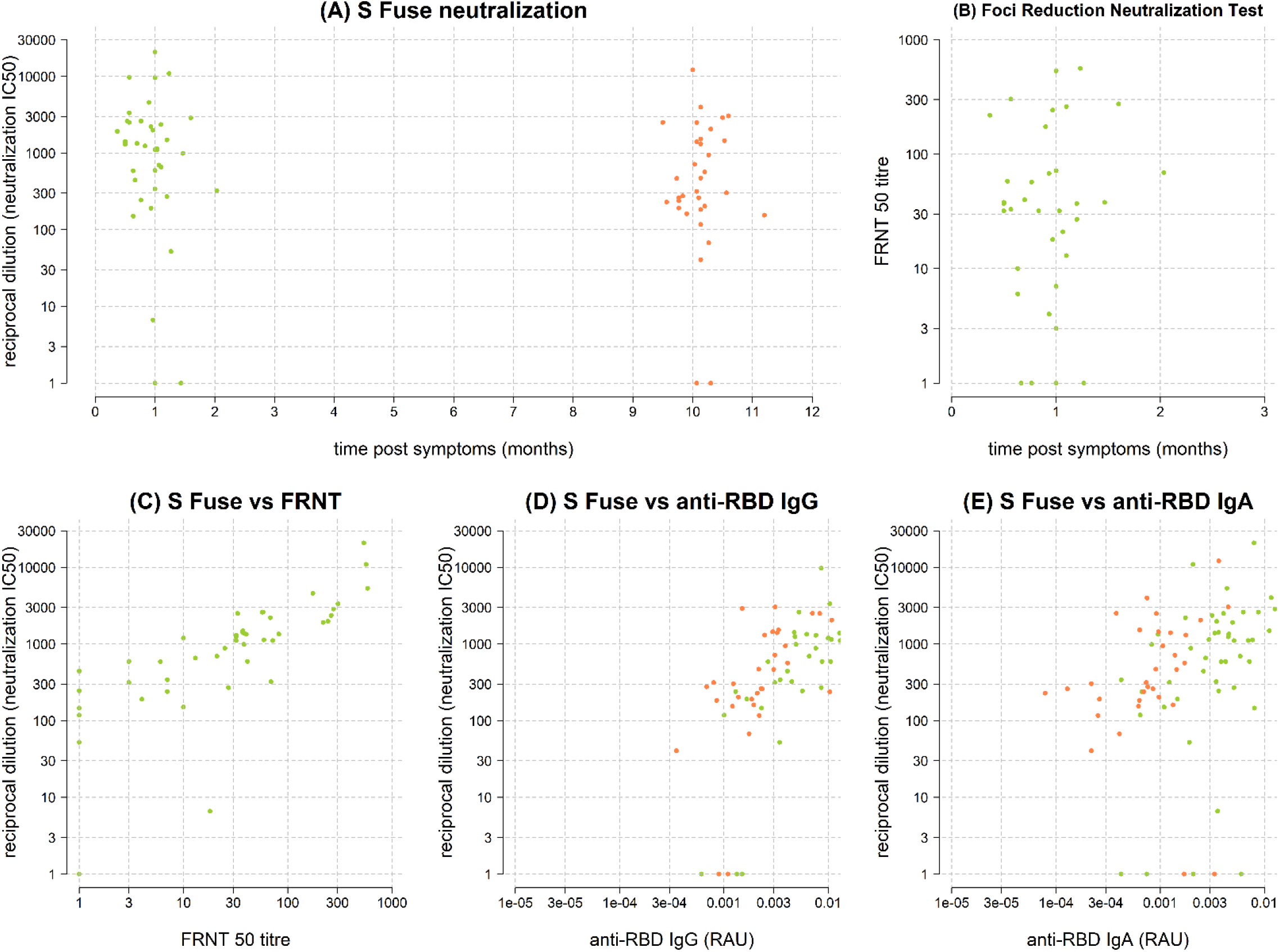
Association between measured antibody levels and viral neutralization. **(A)** Viral neutralization (denoted as reciprocal dilution for 50% neutralization) in serum samples from Institut Mutualiste Montsouris. **(B)** Foci reduction viral neutralization (denoted as reciprocal dilution for 50% neutralization) in serum samples from Institut Mutualiste Montsouris. **(C)** Comparison between two viral neutralization assays. **(D)** Comparison between viral neutralization and anti-RBD IgG. **(E)** Comparison between viral neutralization and anti-RBD IgA.

**Appendix Figure 5:**
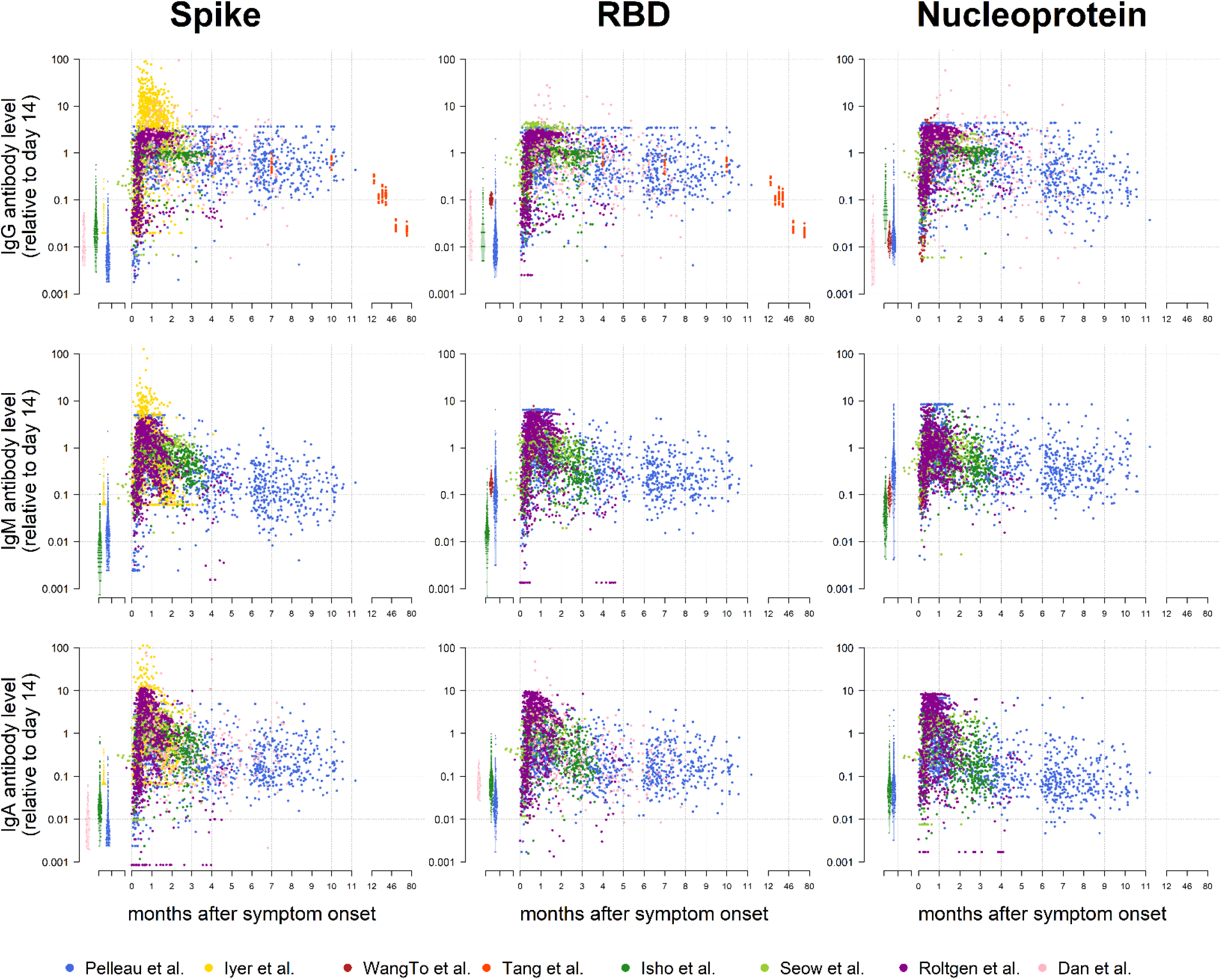
Data on SARS-Cov-2 antibody kinetics. Overview of data used for fitting mathematical model of antibody kinetics from seven studies with longitudinal follow-up of individuals after infection with SARS-CoV-2, and one study with longitudinal follow-up after infection with SARS-CoV-1. Note that the study by Tang *et al*. measured antibodies to SARS-CoV-1 over six years of follow-up, hence the cut in x-axis. There were considerable differences in the immunoassays used for each study, notably variation in antigens and different platforms. For each study, the data has been normalized so that at day 14 post symptom onset, the expected antibody level equals 1. Measured antibody levels from negative controls are shown in the left portion of the plot.

**Appendix Figure 6:**
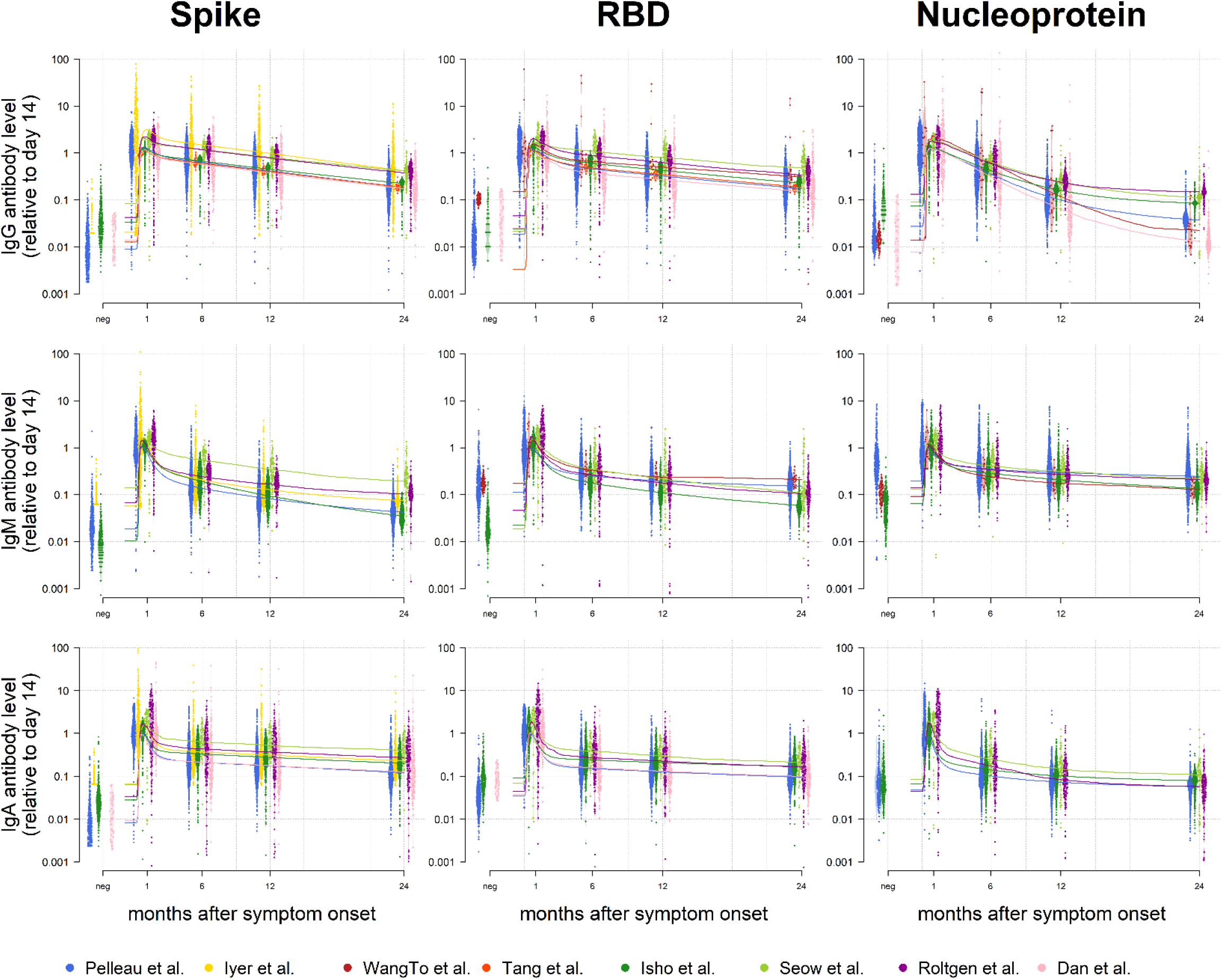
Modelled SARS-CoV-2 antibody kinetics. Overview of the fit of a mathematical model of antibody kinetics to data from seven studies with longitudinal follow-up of individuals after infection with SARS-CoV-2, and one study with longitudinal follow-up after infection with SARS-CoV-1. For each study, the data has been normalized so that at day 14 post symptom onset, the expected antibody level equals 1. Measured antibody levels from negative controls are shown in the left portion of the plot. Predicted antibody kinetics are shown for a period of 2 years. The solid line represents the median predicted antibody level for each study.

**Appendix Figure 7:**
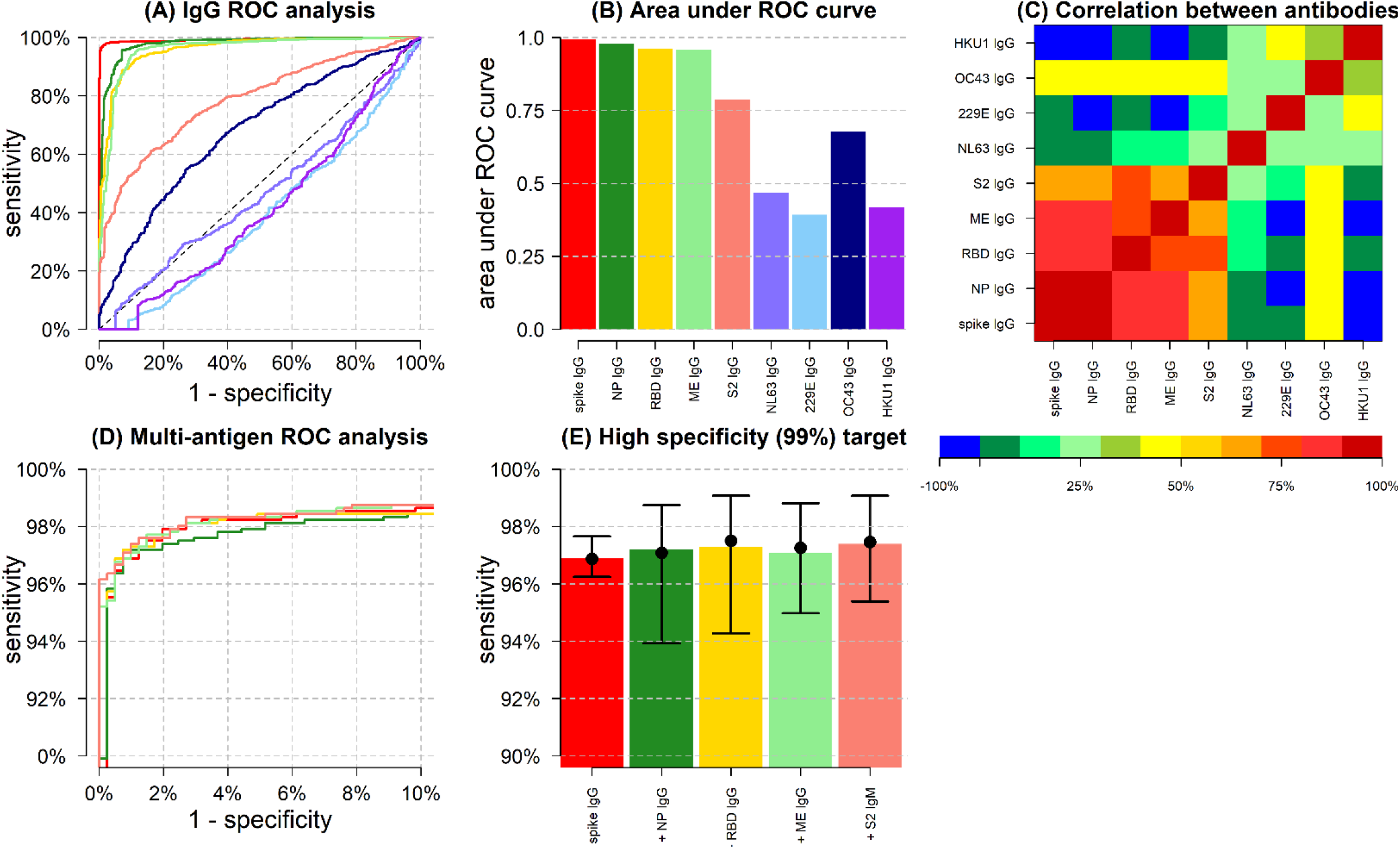
Overview of classification performance of assay and algorithm on IgG data. **(A)** Receiver Operating Characteristic (ROC) curves for IgG antibodies obtained by varying the cutoff for seropositivity. Colours correspond to antibodies against different antigens, as shown in panel B. **(B)** Area under the ROC curve (AUC) for individual IgG antibodies. **(C)** Spearman’s correlation between measured antibody responses. **(D)** ROC curves for multiple biomarker classifiers generated using a random forests algorithm. Biomarkers are added sequentially with colours corresponding to panel E. **(E)** For a high specificity target (>99%), sensitivity increases with additional biomarkers, added sequentially. Sensitivity was estimated using a random forests classifier. Points and whiskers denote the median and 95% CIs from repeat cross-validation.

**Appendix Figure 8:**
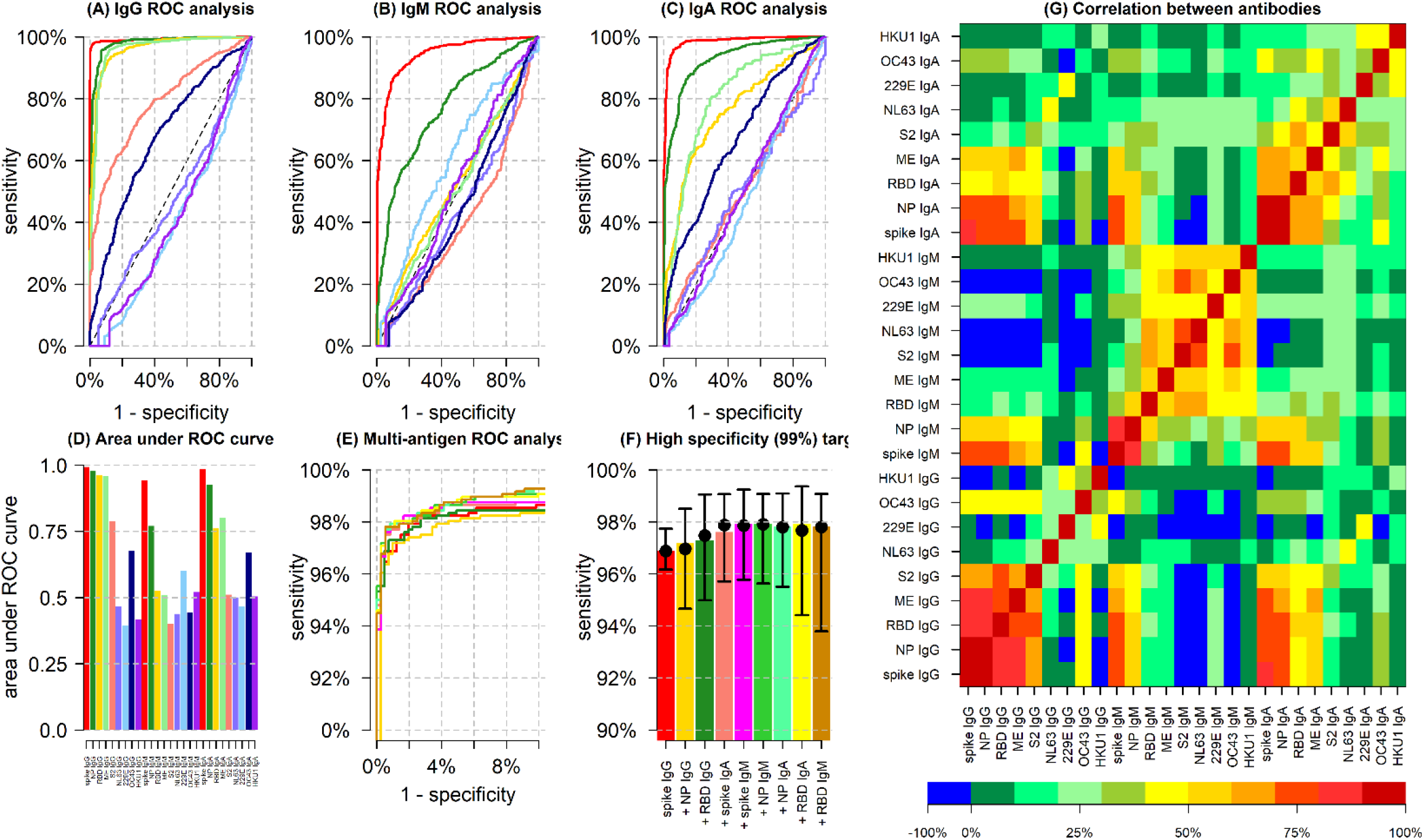
Overview of classification performance of assay and algorithm on IgG, IgM and IgA data. **(A)** Receiver Operating Characteristic (ROC) curves for IgG antibodies obtained by varying the cutoff for seropositivity. Colours correspond to antibodies against different antigens, as shown in panel D. **(B)** ROC curves for IgM antibodies obtained by varying the cutoff for seropositivity. **(C)** ROC curves for IgA antibodies obtained by varying the cutoff for seropositivity. **(D)** Area under the ROC curve (AUC) for individual IgG, IgM and IgA antibodies. **(E)** ROC curves for multiple biomarker classifiers generated using a random forests algorithm. Biomarkers are added sequentially with colours corresponding to panel F. **(F)** For a high specificity target (>99%), sensitivity increases with additional biomarkers, added sequentially. Sensitivity was estimated using a random forests classifier. Points and whiskers denote the median and 95% CIs from repeat cross-validation. **(G)** Spearman’s correlation between measured antibody responses.

## Statistical Appendix

### Data on longitudinal antibody responses to SARS-CoV-2 proteins

Several teams have conducted studies of the duration of the antibody response against SARS-CoV-2 with varying durations of follow up. Our data analyses the duration of the SARS-CoV-2 antibody response over approximately eleven months. To better characterize the antibody kinetics over different time scales, we supplemented our database with data from six published studies on SARS-CoV-2 antibody kinetics. These data are summarized in Appendix Table 1. In addition to the studies on SARS-CoV-2 antibody kinetics, we also included data from one study of SARS-CoV-1 antibody kinetics in individuals with six years follow-up^12^.

**Appendix Table 1:**
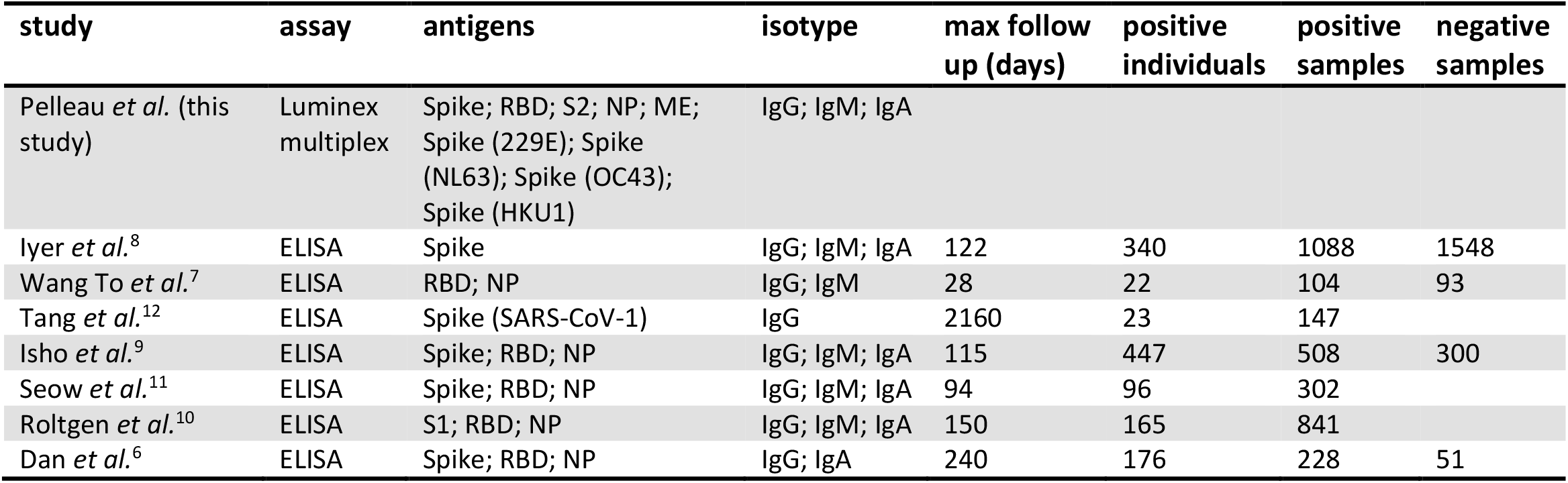
Overview of studies incorporated in antibody kinetic model.

| study | assay | antigens | isotype | max follow up (days) | positive individuals | positive samples | negative samples |
| --- | --- | --- | --- | --- | --- | --- | --- |
| Pelleau *et al.* (this study) | Luminex multiplex | Spike; RBD; S2; NP; ME; Spike (229E); Spike (NL63); Spike (OC43); Spike (HKU1) | IgG; IgM; IgA |  |  |  |  |
| Iyer *et al.*<sup>8</sup> | ELISA | Spike | IgG; IgM; IgA | 122 | 340 | 1088 | 1548 |
| Wang To *et al.*<sup>7</sup> | ELISA | RBD; NP | IgG; IgM | 28 | 22 | 104 | 93 |
| Tang *et al.*<sup>12</sup> | ELISA | Spike (SARS-CoV-1) | IgG | 2160 | 23 | 147 |  |
| Isho *et al.*<sup>9</sup> | ELISA | Spike; RBD; NP | IgG; IgM; IgA | 115 | 447 | 508 | 300 |
| Seow *et al.*<sup>11</sup> | ELISA | Spike; RBD; NP | IgG; IgM; IgA | 94 | 96 | 302 |  |
| Roltgen *et al.*<sup>10</sup> | ELISA | S1; RBD; NP | IgG; IgM; IgA | 150 | 165 | 841 |  |
| Dan *et al.*<sup>6</sup> | ELISA | Spike; RBD; NP | IgG; IgA | 240 | 176 | 228 | 51 |

### Mathematical model of antibody kinetics

SARS-CoV-2 antibody kinetics are described using a previously published mathematical model of the immunological processes underlying the generation and waning of antibody responses following infection or vaccination^20,33^. For antibodies of three classes (IgG, IgM and IgA) targeting the studied antigens, the antibody kinetics are described by the following equations:

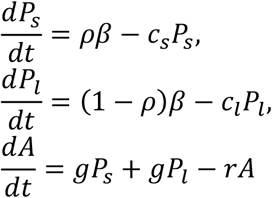

where β denotes the boost in antibody secreting plasma cells. It is assumed that a proportion *ρ* of plasma cells are short-lived (*P*_*s*_) waning at rate *c*_*s*_, and a proportion 1 – *ρ* are long-lived (*P*_*l*_) waning at rate *c*_*l*_. Plasma cells generate antibodies

(IgG, IgM or IgA) at rate *g*, and *r* is the rate of decay of antibody molecules. Assuming *P*_*s*_(0) = *P*_*l*_(0) = 0 and *A*(0) = *A*_*bg*_, these equations can be solved analytically to give:

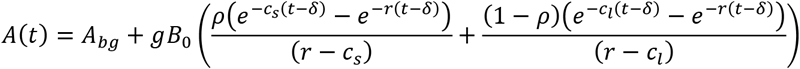

*δ* is the time after symptom onset when antibody levels start to increase. *B*_*0*_ is the number of B cells, and *g* is the rate at which they secrete antibodies. As *g* and *B*_*0*_ are not both identifiable without detailed and invasive experiments (e.g. bone marrow aspirates to measure antigen-specific plasma cells), we estimate *β* = *gB*_0_. If *r* is the decay rate of antibody molecules, then we define 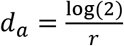 to be the half-life of antibody molecules. Similarly, we define 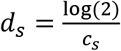 and 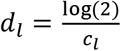.

Note that this model of antibody kinetics follows the formulation outlined in White *et al*.^33^ and not that outlined in Rosado *et al*.^20^ which included an additional equation for modelling the proliferation of memory B cells. Following experiments on model identifiability with simulated data, it was found that there was limited ability to recover model parameters describing memory B cell proliferation.

### Methodology for statistical inference

The model was fitted to longitudinal antibody level measurements from all participants. Mixed effects methods were used to capture the natural variation in antibody kinetics between individual participants, whilst estimating the average value and variance of the immune parameters across the entire population of individuals. The models were fitted in a Bayesian framework using Markov Chain Monte Carlo (MCMC) methods. Mixed effects methods allow individual-level parameters to be estimated for each participant separately, with these individual-level (or mixed effects) parameters being drawn from global distributions.

We consider a scenario where we have data from *M* studies, each of which has data from *N*_*m*_ individuals. For example, for each participant *n* in study *m* the half-life of the short-lived ASCs may be estimated as 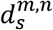 (an individual-level parameter). The 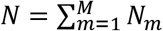 estimates of the local parameters 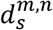 will be drawn from a probability distribution. A log-Normal distribution is suitable as it has positive support on [0, ∞). Thus we have 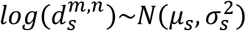. The mean *d*_*s*_ and the variance 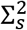 of the estimates of 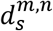 are given by 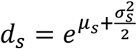 and 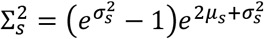.

### Model likelihood

For individual *n* from study *m* we have data on observed antibody levels *A*^*m,n*^= {*a*_1_, …, *a*_*J*_} at times *T*^*m,n*^ = {*t*_1_, …, *t*_*J*_}. We denote *D*^*m,n*^ = (*A*^*m,n*^, *T*^*m,n*^) to be the vector of data for individual *n* from study *m*. For individual *n* from study *m*, the parameters 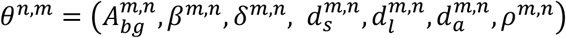 are estimated. The model predicted antibody levels will be {*A*(*t*_1_), *A*(*t*_2_), …, *A*(*t*_*J*_)}. We assume log-Normally distributed measurement error such that the difference between *log*(*a*_*j*_) and *log* (*A*(*t*_*j*_)) is Normally distributed with variance 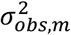. The parameters for observational variance are specific for each study due to differences in the assays utilised. For model predicted antibody levels *A*(*t*_*j*_) the data likelihood for individual *n* from study *m* is given by

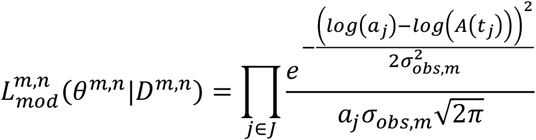

### Mixed effects likelihood

As described above, for each individual there are seven parameters to be estimated. These individual-level parameters are drawn from population-level distributions. Some parameters are assumed not to depend on the assay in each study (the antibody kinetic parameters: 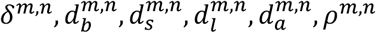). Each of these parameters are assumed to be drawn from the same distribution across all studies. Some parameters are assay dependent, notably 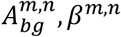. These parameters are drawn from distributions specific for each study. The mixed effects likelihood can be written as follows:

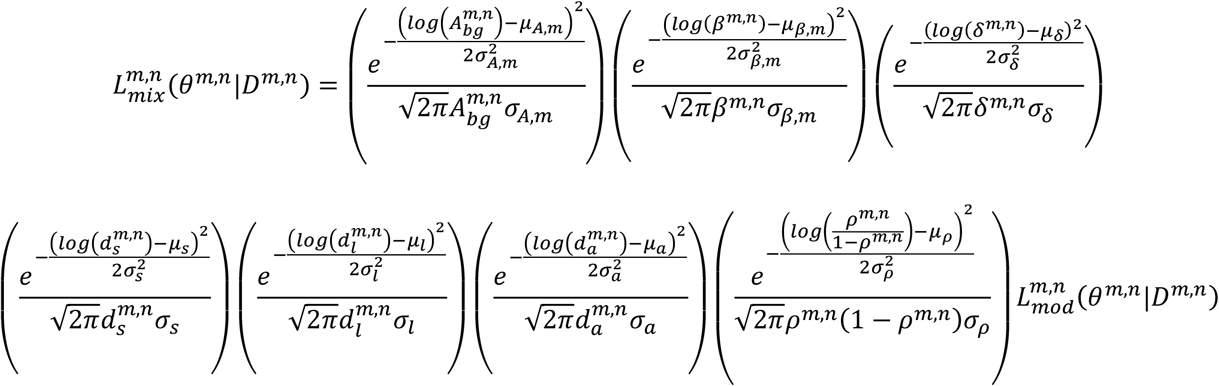

As the proportion of the ASCs that are long-lived must be bounded by 0 and 1, the individual-level parameters *ρ*^*m,n*^ are assumed to be drawn from logit-Normal distributions.

### Total model likelihood

Denote *D* = {*D*^1^, …, *D*^*N*^} to be the vector of data for all *N* participants from all studies. We denote *θ*^*m,n*^ = (*μ*_*A,m*_, *σ*_*A,m*_, *μ*_*β,m*_, *σ*_*β,m*_, *μ*_*δ*_, *σ*_*δ*_, *μ*_*s*_, *σ*_*s*_, *μ*_𝓁_, *σ*_𝓁_, *μ*_a_, *σ*_*a*_, *μ*_*ρ*_, *σ*_*ρ*_, *θ*^1^, …, *θ*^*N*^) to be the combined vector of population-level parameters and individual-level parameters to be estimated. The total likelihood is obtained by multiplying the likelihood for each participant

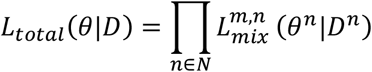

### Markov Chain Monte Carlo parameter update

The model was fitted to the data using Markov Chain Monte Carlo (MCMC) methods. A three stage parameter update regimen was utilised with a Metropolis-within-Gibbs sampler with sequential updating of individual-level parameters, population-level parameters, and observational variance parameters. A′ indicates an attempted update.

#### 1. Individual-level parameter Metropolis-Hastings update

For each participant *n* in study *m*:

- Update local parameters: 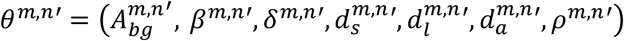
- Calculate updated mixed effects likelihood 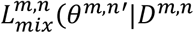
- Accept the parameter update with probability 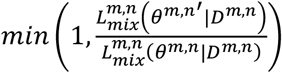

#### 2. Population-level parameter Gibbs update

- For each of the 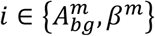 we obtain new estimates of the population level parameters *μ*_*i*_′ and 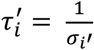 as follows:

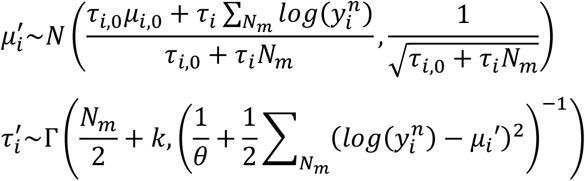

where *μ*_*i*,0_ and τ_*i*,0_ parameterise the Normal prior distribution on the mean, and *k* and θ parameterise the Gamma prior distribution on the precision (inverse standard deviation). 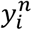 is the individual-level parameter *i* in individual *n*.
- For each of the *i* ∊ {*δ, s*, 𝓁, *a*} we obtain new estimates of the population level parameters *μ*_*i*_′ and 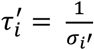 as follows:

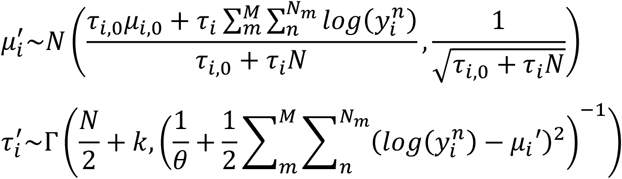

where *μ*_*i*,0_ and τ_*i*,0_ parameterise the Normal prior distribution on the mean, and *k* and θ parameterise the Gamma prior distribution on the precision (inverse standard deviation). 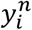 is the individual-level parameter *i* in individual *n*.
- For *ρ* we obtain new estimates of the population level parameters *μ* ′ and 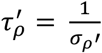 as follows:

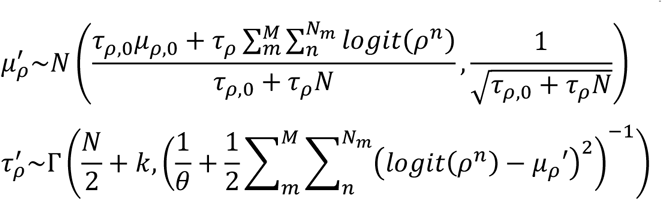

where *μ*_*ρ*,0_ and τ_*ρ*,0_ parameterise the Normal prior distribution on the mean, and *k* and θ parameterise the Gamma prior distribution on the precision (inverse standard deviation).

#### 3. Observational variance parameter Metropolis-Hastings Update

- For each study *m*, update the observational variance parameter *σ*_*ob*s,*m*_′
- Calculate updated total likelihood *L*_*tot*a𝓁_(*θ*′|*D*)and the updated prior probability density *P*(*θ*′)
- Accept the parameter update with probability 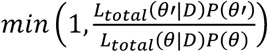

The MCMC algorithm was implemented in C++ complied in Microsoft Visual Studio. The covariance of the multivariate-Normal proposal distributions for Metropolis-Hastings updates were adaptively tuned using the estimated posterior distributions during a burn-in phase of 1 million MCMC iterations. The magnitude of the proposed step size was calibrated using a Robbins-Munro algorithm to achieve an acceptance rate of approximately 23%. The total number of MCMC iterations was 10,000,000. The effective number of iterations was calculated using the effectiveSize routine in the R library coda and the effective size was checked to be > 1,000 for all parameters. The statistical inference procedure was repeated twice to allow for assessment of chain convergence using Gelman-Rubin convergence diagnostics *R*_*c*_. For all population-level chains, we ensure *R*_*c*_< 1.05. For all individual-level chains, we ensure *R*_*c*_ < 1.1.

### Serological surveillance algorithms

A ROC curve obtained from a training data set consisting of positive and negative samples is described by a sequence of estimated sensitivities and specificities{*E*(*se*_*i*_, *sp*_*i*_)}, where *E* denotes an estimator. *N*-fold cross-validation generates samples of sensitivity {s*e*_*i*1_, …, *se*_*iN*_} for each *E*(s*p*_*i*_) and samples of specificity {*sp*_*i*1_, …, *sp*_*iN*_} for each E(*se*_*i*_). Following a previously outlined approach^34^, for each pair *i* of sensitivity and specificity, we obtain *N* estimates of the measured seroprevalence *M*_*in*_ in a scenario with true seroprevalence *T* as follows:

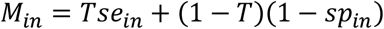

Following the Rogan-Gladen estimator approach, this equation can be rearranged to give an adjusted estimate of true seroprevalence:

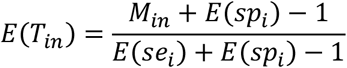

with *E*(*T*_*in*_) = 0 if *M*_*in*_ < 1 − *E*(s*e*_*i*_). Both *M*_*i*_ and *E*(*T*_*i*_) are summarized as medians with 95% ranges. Define *α*^*K*^ as a *K*-way classification algorithm that can classify a sample into one of *K* categories. Let *C*_0_ be the category for a sample from an individual not previously infected with SARS-CoV-2. Let *C*_*k*_ be the category for individuals previously infected with SARS-CoV-2 in one of *K* – 1 time intervals defined by (0, *t*_*1*_, …, *t*_*K*-1_). *α*^*K*^ can be any of a range of algorithms from Random Forests, Support Vector Machines, or ordinal logistic regression. *α*^*K*^ can be trained on a data set, with classification performance assessed using *N*-fold cross-validation. For a serum sample, let *P*_*k*_ denote the classification score (analogous to a probability) that this sample belongs to category *k*.

Given a vector of classification scores *P*_*k*_, we can assign a serum sample to a category. A frequently used approach is to select *k* according to the maximum of *P*_*k*_. In order to retain better control over diagnostic specificity, we select an alternative two step approach:

- if *P*_0_ ≥ γ_99_ then the sample is classified as negative
- if *P*_*0*_ *<* γ_99_ then the sample is classified into category *k* according to the maximum of *P*_*1*_, …, *P*_*K*-1_

Here γ_99_ is selected based on the training data to ensure a target specificity of 99%. The classification performance of this approach can be assessed using a confusion matrix *C*_*ij*_ defined by the proportion of samples of category *i* classified as category *j*. For the example with *K* = 4 explored in more detail here, we define three intervals: 0 – 3 months; 3 – 6 months; 6 – 12 months.

Following the approach of the Rogan-Gladen estimator, define ***M*** to be the vector of measured prevalences in each of the K categories. An estimator of the true sero-prevalence is provided by E(***T***) = ***C***^-1^ * ***M*** where ***C***^-1^ is the inverse of the confusion matrix. Depending on the values of the confusion matrix, this approach may have *T*_*k*_ < 0. In this instance we assign *T*_*k*_ = 0 and transfer the predictions proportionally to the other categories.

